# Specific associations between plasma biomarkers and post-mortem amyloid plaque and neurofibrillary tau tangle loads

**DOI:** 10.1101/2022.08.22.22279052

**Authors:** Gemma Salvadó, Rik Ossenkoppele, Nicholas J. Ashton, Thomas G. Beach, Geidy E. Serrano, Henrik Zetterberg, Niklas Mattsson-Carlgren, Shorena Janelidze, Kaj Blennow, Oskar Hansson

## Abstract

Several promising plasma biomarkers have recently been developed that could serve as diagnostic and/or prognostic tools for Alzheimer’s disease (AD). However, their neuropathological correlates have not yet been fully determined. Therefore, we aimed to investigate the independent associations between multiple plasma biomarkers (*i*.*e*., phosphorylated tau217 [p-tau217], p-tau181, p-tau231, the amyloid-β_42/40_ [Aβ42/40] ratio, glial fibrillary acidic protein [GFAP] and neurofilament light [NfL]) and core semi-quantitative measures of AD pathology (*i*.*e*., amyloid plaques and tau neurofibrillary tangles) as well as common co-pathologies (*i*.*e*., cerebral amyloid angiopathy, Lewy body disease, TAR DNA-binding protein 43, cerebral white matter rarefaction and argyrophilic grain disease). We included 105 participants from the Arizona Study of Aging and Neurodegenerative Disorders and Brain and Body Donation Program with antemortem collected plasma samples and a post-mortem neuropathological exam (mean(SD) time: 482(355) days), 48 of whom had longitudinal p-tau217 and p-tau181 (mean(SD) follow-up time: 1,378(1,357) days). Participants ranged from cognitively unimpaired to Alzheimer’s and non-Alzheimer’s dementia. All markers except NfL were associated with plaques (| β|≥0.37, p<0.001) and tangles (| β|≥0.27, p<0.008), in univariable analyses adjusted for age, sex and time between blood sampling and death. In multivariable models, when including both plaques and tangles as independent variables, the Aβ42/40 ratio and p-tau231 were only associated with plaques (β_Aβ42/40_ [95%CI]=-0.59[-0.80,-0.38], R^2^_plaques_/R^2^=77.6%; β_p-tau231_[95%CI]=0.32[0.09,0.56], R^2^_plaques_/R^2^=45.9%, all p≤0.007), while GFAP was only associated with tangles (β_GFAP_[95%CI]=0.39[0.19,0.59], p<0.001, R^2^_tangles_/R^2^=30.4%). In contrast, p-tau217 and p-tau181 were associated with both plaques (β_p-tau217_[95%CI]=0.46[0.30,0.62], R^2^_plaques_/R^2^=40.4%; β_p-tau181_[95%CI]=0.41[0.22,0.60], R^2^_plaques_/R^2^=35.7%, both p<0.001) and tangles (β_p-tau217_[95%CI]=0.40[0.24,0.57], p<0.001, R^2^_tangles_/R^2^=30.7%; β_p-tau181_[95%CI]=0.30[0.10,0.49], p=0.004, R^2^_tangles_/R^2^=17.1%). A parsimonious model predicting plaque load included p-tau217 and Aβ42/40, while a parsimonious model for tangle burden included only p-tau217. Further, combining p-tau217 and Aβ42/40 ratio yielded the highest accuracy for predicting intermediate/high AD neuropathological change ([ADNC], AUC[95%CI]=0.90[0.84,0.96],R^2^=0.66). High plasma NfL levels were predictive of presence of cerebral white matter rarefaction (AUC[95%CI]=0.76[0.66,0.85],R^2^=0.25). Finally, p-tau217 (β[95%CI]=0.13[0.02,0.24], p=0.018), but not p-tau181 (β[95%CI]=0.12[-0.05,0.29], p=0.152), levels increased more over time in participants with intermediate/high ADNC compared with those with none/low ADNC. In this relatively large neuropathological study with multiple plasma biomarkers available, we showed that the Aβ42/40 ratio and p-tau231 were specific markers of plaque pathology, and GFAP of tangle pathology, while p-tau181 and, especially, p-tau217 were markers of both plaque and tangle pathologies. Our results suggest that high-performing assays of plasma p-tau217 and Aβ42/40 might be an optimal biomarker combination to detect ADNC *in vivo*.

## Introduction

The recent development of plasma biomarkers for Alzheimer’s disease (AD) has revolutionized the field,^1–3^ as these markers have the benefit of being significantly cheaper and less invasive than established markers (*i*.*e*., cerebrospinal fluid [CSF] and positron emission tomography [PET]), while showing excellent diagnostic performance.^4–6^ Several plasma biomarkers are currently available, among which the most studied include the amyloid-β_42/40_ (Aβ42/40) ratio, glial fibrillary acidic protein (GFAP), neurofilament light (NfL), and, particularly, phosphorylated tau (p-tau) measures. Previous studies have indicated excellent diagnostic performance of some of these plasma biomarkers for distinguishing AD from non-AD neurodegenerative disorders,^4,6–9^ with non-inferior performance compared with CSF and PET markers ^5,9–13^ as well as important utility for predicting disease progression.^5,11,13,14^ Nonetheless, there are still important topics to be addressed to optimize their usage in clinical practice, including improved interpretation of obtained plasma biomarker results and a fair head-to-head comparison, especially against gold standard neuropathological measures.^15^

One of the most important knowledge voids of plasma biomarkers is the degree to which they specifically correlate with key neuropathological changes. Although previous studies have already investigated the association of some of these biomarkers with measures of neuropathology, whether these markers are primarily related to amyloid, tau, or to both pathologies is still under debate. For instance, p-tau181, has shown strong associations with neuropathological measures of amyloid-β and tau pathologies,^8,16–18^ but this has been shown in independent analyses for amyloid and tau or with scales combining these two pathologies, which did not allow for the interpretation of the specific -or independent-associations with these two pathological measures. Similarly, plasma p-tau231 has also shown associations with neuropathologically defined plaque and tangle load, without exploring specific associations with these neuropathologic measures.^13,17^ Only one study with plasma p-tau217 has suggested that this biomarker may be independently associated with both plaques and tangles.^19^ On the other hand, the accuracy of the plasma Aβ42/40 ratio, GFAP, or NfL levels to predict AD pathology seems to be lower than that of p-tau markers, although only few studies have investigated their association with neuropathologic measures of AD pathology.^8,17,20^

Another challenge when trying to optimize the use of plasma biomarkers in clinical practice is the lack of comparison among biomarkers in the same population. Differences in clinical performance for the same biomarkers can be observed depending on the characteristics of the study (*e*.*g*., diagnostic groups, patient characteristics, outcomes and/or presence of co-pathologies). Thus, head-to-head studies are crucial to allow a fair comparison and avoid bias due to population selection. Nonetheless these studies are scarce, especially those including neuropathological measures.^17,20^ This would also allow investigating whether any of these biomarkers might be useful for detecting other common co-pathologies observed in AD patients, such as Lewy bodies or TAR DNA-binding protein 43 (TDP-43).^1,17^

When comparing multiple plasma biomarkers, it is equally important to consider the discriminative power of the assays. While there are many plasma biomarkers currently available, there are also many platforms by which to measure them, which can highly affect their performance. As it has been shown recently with the plasma Aβ42/40 ratio, different assays and/or platforms could lead to significantly different performances in detecting AD-related pathology.^21^ Similarly, previous comparisons between multiple species and assays of p-tau measures showed only a modest correlation, suggesting also significantly different diagnostic performance.^11,22^ Considering the differences in clinical performance of different assays for the same biomarkers, importance the use of high-performing assays is of utmost when comparing different biomarkers to avoid reporting differences that are related to the method rather than the biomarkers themselves.

Therefore, the main objective of this study was to identify specific relationships between multiple plasma biomarkers and core AD-related pathologies using high performing assays. To this end, we investigated associations between multiple plasma biomarkers and autopsy-assessed measures of core AD pathologies (plaque and tangle load) in the same participants. We focused on investigating whether these biomarkers primarily reflect amyloid, tau or both pathologies. Further, we identified the best combination of biomarkers to predict each of these pathological measures as well as the presence or absence of AD as a binary measure based on pathology.^23^ We also investigated associations between plasma biomarkers and presence of co-pathologies commonly observed in AD patients including cerebral amyloid angiopathy (CAA), Lewy body disease (LBD), TDP-43, cerebral white matter rarefaction (CWMR) and argyrophilic grain disease (AGD). Finally, we examined whether longitudinal changes of the two plasma biomarkers longitudinally available (*i*.*e*., p-tau217 and p-tau181) were associated with presence of AD pathology.

## Materials and methods

### Participants

All samples were obtained through autopsies of subjects enrolled in the Arizona Study of Aging and Neurodegenerative Disorders and Brain and Body Donation Program (BBDP) at Banner Sun Health Research Institute.^24^ The BBDP recruits independently-living normal and neurologically-impaired elderly subjects predominantly from the surrounding Sun City’s retirement communities. These volunteer research subjects are followed prospectively with annual standardized clinical assessments for the rest of their lives. Participants included in this study ranged from cognitively unimpaired to mild cognitive impairment and AD patients as well as patients with other neurodegenerative diseases. We selected participants with both plasma and neuropathological exam available, including only those with all biomarkers available in the cross-sectional analyses. Participants in the cross-sectional analysis were also restricted as to those having blood drawing up to five years before death (mean(SD) [range] time: 482 (355) [9 - 1760] days). All experiments were conducted in accordance with the Declaration of Helsinki. The operations of the Brain and Body Donation Program are approved by Institutional Review Boards and all participants or their legal representatives gave informed consent.

### Plasma biomarkers

Plasma p-tau217 and p-tau181 concentrations were measured in-house using an immunoassay developed by Lilly Research Laboratories (IN, USA), each of which had performed very well in multiple studies and cohorts.^6,9,11,25^ Plasma p-tau231 concentration was also measured in-house using a Simoa approach which was developed at the University of Gothenburg, which can detect Aβ pathology with high accuracy.^13^ The remaining plasma biomarkers (Aβ42, Aβ40, GFAP and NfL) were measured with prototype fully automated Elecsys^®^ plasma immunoassays (updated versions for Aβ42 and Aβ40 ^26^) plasma immunoassays (not commercially available) cobas e 601 and cobas e 411 analyzers (Roche Diagnostics International Ltd, Rotkreuz, Switzerland), also in-house.^12^ Longitudinal samples were only analysed for p-tau217 and p-tau181, and not the Elecsys measurements or p-tau231.

Outliers were defined as subjects with values above or below more than 5 interquartile range of the third or the first quartile and were excluded from subsequent analyses. Only one plasma NfL value was considered an outlier. We also excluded another plasma NfL value that strongly affected the models (based on the standardized residuals) due to a very young age of the subject (44 years) and the strong correlation between NfL and age.

### Pathological measures

#### Core AD pathology

Tissue processing methods have been detailed previously.^27^ Histopathological scoring was performed blinded to clinical and neuropathological diagnosis as well as levels of the plasma biomarkers. Amyloid plaque and neurofibrillary tangle density were graded at standard sites in frontal, temporal, parietal, and occipital cortices as well as hippocampus and entorhinal cortex, as previously explained.^27^ To obtain the total plaque score, each region was first rated as none, sparse, moderate, or frequent, using the published Consortium to Establish a Registry for Alzheimer’s Disease (CERAD) templates.^28^ These descriptive measures were then converted into 0-3 scores in each region that combined, gave the total plaque score with a maximum value of 15. Neurofibrillary tangle abundance was measured similarly using the CERAD templates. Additionally, Braak staging was performed based on the topographical distribution of neurofibrillary tangle change.^29^ Global CERAD scoring and Thal phases^30^ were also assessed as a measure of amyloid pathology. Using these three global scales we obtained a global measure of AD neuropathologic change (ADNC) as described in the NIA-AA guidelines.^23^ Although the ADNC score was used as a four-scale measure in some analyses, we also dichotomized as a positive (negative) ADNC when scores were intermediate or high (none or low).

#### Non-AD pathology

CAA was graded on a 0-3 scale based analogously on CERAD templates^28^ and dichotomized as positive if the score was above 1. Immunohistochemical staining in 10 brain regions for p129 alpha-synuclein, as well as Thioflavin-S (for substantia nigra) was used as a secondary stain to detect Lewy bodies (LBs) and were staged based on the Unified Staging System.^31^ TDP-43 positivity, location of positivity, and morphology were recorded as explained previously.^32^ Significant CWMR was defined as exceeding 25% of the total centrum semiovale area within one or more cerebral lobes using hematoxylin and eosin on large format section protocol.^33^ AGD was defined as typical spindle-shaped structures revealed by the Gallyas silver stain.^34,35^

### Statistical analyses

First, linear regression models were used to assess associations between each plasma biomarker (as dependent variable) and both plaques and tangles (as independent variables), independently. These models were adjusted for age, sex and time between blood draw and death. To assess specific associations with each of these pathological measures, we performed multivariable linear regression models with plasma biomarkers as outcomes and measures of both plaques and tangles as predictors using the same covariates. Differences between two correlation coefficients were tested using a bootstrapping approach as described in Rosner et al.^36^ Specific contribution of plaque and tangle loads on each biomarker concentration was obtained as the percentage of partial R^2^ of each pathology over the total R^2^ of the multivariable model. Diagnostic accuracies of plasma biomarkers were assessed using receiver operating characteristic (ROC) curve analysis, with age sex and time between blood draw and death as covariates. When assessing diagnostic accuracy of non-AD pathologies, the ADNC status as a dichotomous variable was also included as a covariate. Differences in area under the curve (AUC) between two ROC curves were compared with the DeLong test.^37^ Differences in plasma levels between pathological groups were assessed with Kruskal-Wallis tests, with pairwise Wilcoxon test as a *post hoc* comparison among groups (only differences between consecutive groups tested). We used the R package MuMIn to select the most parsimonious models to predict both continuous and dichotomous pathological measures following procedures described earlier.^38^ Only plasma biomarkers showing a significant association (p<0.05) with each pathological measure in the initial univariable models were included as possible predictors as well as the aforementioned covariates. All covariates were included in the final parsimonious model (even when they were not significant) to allow a fair comparison against univariable models. When multiple p-tau (at different phosphorylation sites) biomarkers were possible predictors, independent models for each p-tau marker were performed and then compared based on the corrected Akaike criterion (AICc) to avoid multicollinearity problems. Linear mixed models (LME) were used to assess longitudinal changes on plasma biomarkers. Three independent models were performed for each biomarker. In each one, the interaction between time and one measure of AD pathology (i. plaques, ii. tangles or iii. presence of ADNC) was used as predictor. The LME models were adjusted for age and sex and included random intercepts and fixed slopes due to the limited number of datapoints (median[range]: 2[2-5]). Log-transformed biomarker measures were used in regression analyses. Statistical analyses were done using R version 4.1.0. Significance was set at p<0.05 (two-tailed).

### Data availability

Anonymized data will be shared by request from a qualified academic investigator for the sole purpose of replicating procedures and results presented in the article.

## Results

We included 46 participants with none/low ADNC and 59 participants with intermediate/high ADNC (Table 1). No differences in age at death nor sex were observed between groups. *APOE-ε4* prevalence (49.2% *vs*. 10.9%, p<0.001) and core AD pathology (plaques: 12.70 *vs*. 1.03, p<0.001; tangles: 9.79 *vs*. 5.54, p<0.001) measures were significantly higher in participants with intermediate/high ADNC. Among all the co-pathologies under investigation, only the presence of CAA was higher in participants with intermediate/high ADNC (86.4% *vs*. 34.8%, p<0.001).

**Table 1.** Demographic characteristics at baseline. ADNC was dichotomized as: negative (none/low) and positive (intermediate/high). ^#^ Forty-seven participants missing * One participant missing Abbreviations: A β, amyloid-β; ADNC, Alzheimer’s disease neuropathologic change; AGD, argyrophilic grains disease; CAA, cerebral amyloid angiopathy; CERAD, Consortium to Establish a Registry for Alzheimer’s Disease; CWMR, cerebral white matter rarefaction; GFAP, glial fibrillary acidic protein; LBD, Lewy body disease; NfL, neurofilament light; p-tau, phosphorylated tau; TDP-43, TAR DNA binding protein 43.

|  | Overall<br>(n=105) | ADNC –<br>negative<br>(n=46) | ADNC –<br>positive<br>(n=59) | p-value |
| --- | --- | --- | --- | --- |
| **Age, mean(SD)** | 84.7 (7.99) | 83.5 (7.99) | 85.7 (7.93) | 0.174 |
| **Women, n(%)** | 43 (41.0%) | 21 (45.7%) | 22 (37.3%) | 0.506 |
| **APOE-e4 carrier, n (%)** | 34 (32.4%) | 5 (10.9%) | 29 (49.2%) | <0.001 |
| **Time between blood and**<br>**death, days, mean(SD) [range]** | 482 (355)<br>[9 - 1760] | 414 (300)<br>[9 - 1120] | 536 (387)<br>[9 - 1760] | 0.137 |
| **Plaque total, mean(SD)** | 7.60 (6.34) | 1.03 (1.99) | 12.7 (2.83) | <0.001 |
| **CERAD moderate/frequent,**<br>**n(%)** | 59 (56.2%) | 1 (2.2%) | 58 (98.3%) | <0.001 |
| **Tangle total, mean(SD)** | 7.93 (3.69) | 5.54 (2.41) | 9.79 (3.44) | <0.001 |
| **Braak stage, n(%)** |  |  |  |  |
| **I-II** | 5 (4.8%) | 5 (10.9%) | 0 (0%) | <0.001 |
| **III-IV** | 65 (61.9%) | 37 (80.4%) | 28 (47.5%) |  |
| **V-VI** | 35 (33.3%) | 4 (8.7%) | 31 (52.5%) |  |
| **CAA, n(%)** | 67 (63.8%) | 16 (34.8%) | 51 (86.4%) | <0.001 |
| **LBD, n(%)** | 21 (20.0%) | 11 (23.9%) | 10 (16.9%) | 0.523 |
| **TDP-43, n(%)<sup>#</sup>** | 19 (18.1%) | 5 (10.9%) | 14 (23.7%) | 0.422 |
| **CWMR, n(%)** | 58 (55.2%) | 20 (43.5%) | 38 (64.4%) | 0.052 |
| **AGD, n(%)<sup>\*</sup>** | 28 (26.7%) | 12 (26.1%) | 16 (27.1%) | 1.000 |
| **p-tau217, pg/ml** | 0.432<br>(0.394) | 0.194<br>(0.113) | 0.617<br>(0.434) | <0.001 |
| **p-tau181, pg/ml** | 2.36 (1.49) | 1.61 (0.845) | 2.96 (1.61) | <0.001 |
| **p-tau231, pg/ml** | 30.0 (15.2) | 25.4 (13.6) | 33.6 (15.5) | <0.001 |
| **Aβ42/40** | 0.125<br>(0.0142) | 0.133<br>(0.0135) | 0.119<br>(0.0114) | <0.001 |
| **GFAP, ng/ml** | 0.190<br>(0.110) | 0.149<br>(0.0948) | 0.222<br>(0.111) | <0.001 |
| **NfL, pg/ml** | 7.37 (5.11) | 7.17 (5.12) | 7.53 (5.15) | 0.420 |

### Associations between plasma biomarkers and core AD pathologies

First, we studied possible associations between plasma biomarkers and total amount of plaques and tangles, independently. All plasma biomarkers except NfL (β=0.13, p=0.166) were significantly associated with the total amount of plaques (0.37≤| β|≤0.69, p<0.001, Figure 1 and Table 2). Plasma p-tau217 showed the highest correlation coefficient with plaques, which was significantly higher than all others (0.11≤ β_diff_≤0.56, p≤0.014) except plasma Aβ42/40 ratio (β_diff_= 0.15, p=0.090). All plasma biomarkers except NfL (β=0.18, p=0.053) were also associated with the total amount of tangles in the independent models (0.27≤| β|≤0.69, p≤0.008; Figure 1 and Table 2). Again, p-tau217 had the highest correlation coefficient with total amount of tangles, which was significantly higher than all other plasma correlation coefficients (0.18≤ β_diff_≤0.51, p≤0.037).

**Table 2.** Associations between plasma biomarkers and amyloid plaque and/or neurofibrillary tau tangle loads. Linear regression models were used to investigate these associations in independent models including: age, sex, and time between blood sampling and death as covariates. Semi-quantitative amyloid plaque load was used as predictor in the first set of analyses. In the second set, semi-quantitative tau tangle load was used as predictor. Finally, both amyloid plaque load and tau tangle load were included in all models as predictors, in the last set of analyses. Significant associations (p<0.05) are shown in bold. Differences between the correlation coefficients were tested using estimated standardized betas (β) coefficients and the method described in Rosner et al. ^36^, using the strongest association as reference (ref.) and shown in the last column. Significant differences (p<0.05) can be understood as significantly weaker associations compared with those of the references (ref.) in each case. Abbreviations: A β amyloid-β; AICc, corrected Akaike criterion, CI, confidence intervals; GFAP, glial fibrillary acidic protein; NfL, neurofilament light; p-tau, phosphorylated tau.

| | $\beta$ plaques<br>[95%CI] | p-value<br>plaques<br>$\beta$ | p-value<br>strengt<br>h<br>plaques | $\beta$ tangles<br>[95%CI] | p-value<br>tangles $\beta$ | p-value<br>strengt<br>h<br>tangles | R<sup>2</sup> | AICc |
| --- | --- | --- | --- | --- | --- | --- | --- | --- |
| **Plaques** |  |  |  |  |  |  |  |  |
| p-tau217 | 0.69<br>[0.55, 0.83] | <0.001 | ref. | - | - | - | 0.50 | 233.82 |
| p-tau181 | 0.58<br>[0.43, 0.74] | <0.001 | 0.014 | - | - | - | 0.38 | 255.52 |
| p-tau231 | 0.37<br>[0.19, 0.56] | <0.001 | 0.001 | - | - | - | 0.14 | 289.75 |
| Aβ42/40 | -0.53<br>[-0.70, -0.37] | <0.001 | 0.090 | - | - | - | 0.30 | 267.61 |
| GFAP | 0.41<br>[0.25, 0.57] | <0.001 | 0.001 | - | - | - | 0.33 | 264.12 |
| NfL | 0.13<br>[-0.05, 0.30] | 0.166 | <0.001 | - | - | - | 0.20 | 281.64 |
| **Tangles** |  |  |  |  |  |  |  |  |
| p-tau217 | - | - | - | 0.69<br>[0.54, 0.85] | <0.001 | ref. | 0.46 | 241.41 |
| p-tau181 | - | - | - | 0.56<br>[0.38, 0.73] | <0.001 | 0.005 | 0.32 | 264.85 |
| p-tau231 | - | - | - | 0.29<br>[0.09, 0.49] | 0.005 | <0.001 | 0.08 | 297.05 |
| Aβ42/40 | - | - | - | -0.27<br>[-0.47, -0.07] | 0.008 | <0.001 | 0.09 | 295.96 |
| GFAP | - | - | - | 0.51<br>[0.35, 0.67] | <0.001 | 0.037 | 0.40 | 252.87 |
| NfL | - | - | - | 0.18<br>[0.00, 0.37] | 0.053 | <0.001 | 0.22 | 279.72 |
| **Plaques and Tangles** |  |  |  |  |  |  |  |  |
| p-tau217 | 0.46<br>[0.30, 0.62] | <0.001 | 0.163 | 0.40<br>[0.24, 0.57] | <0.001 | Ref. | 0.59 | 214.24 |
| p-tau181 | 0.41<br>[0.22, 0.60] | <0.001 | 0.114 | 0.30<br>[0.10, 0.49] | 0.004 | 0.052 | 0.43 | 248.83 |
| p-tau231 | 0.32<br>[0.09, 0.56] | 0.007 | 0.022 | 0.09<br>[-0.15, 0.33] | 0.466 | 0.002 | 0.14 | 291.48 |
| Aβ42/40 | -0.59<br>[-0.80, -0.38] | <0.001 | Ref. | 0.10<br>[-0.12, 0.32] | 0.374 | <0.001 | 0.30 | 269.07 |
| GFAP | 0.19<br>[-0.01, 0.38] | 0.058 | 0.008 | 0.39<br>[0.19, 0.59] | <0.001 | 0.938 | 0.41 | 251.33 |
| NfL | 0.03<br>[-0.19, 0.25] | 0.771 | <0.001 | 0.16<br>[-0.07, 0.39] | 0.169 | 0.078 | 0.21 | 281.93 |

**Figure 1.**
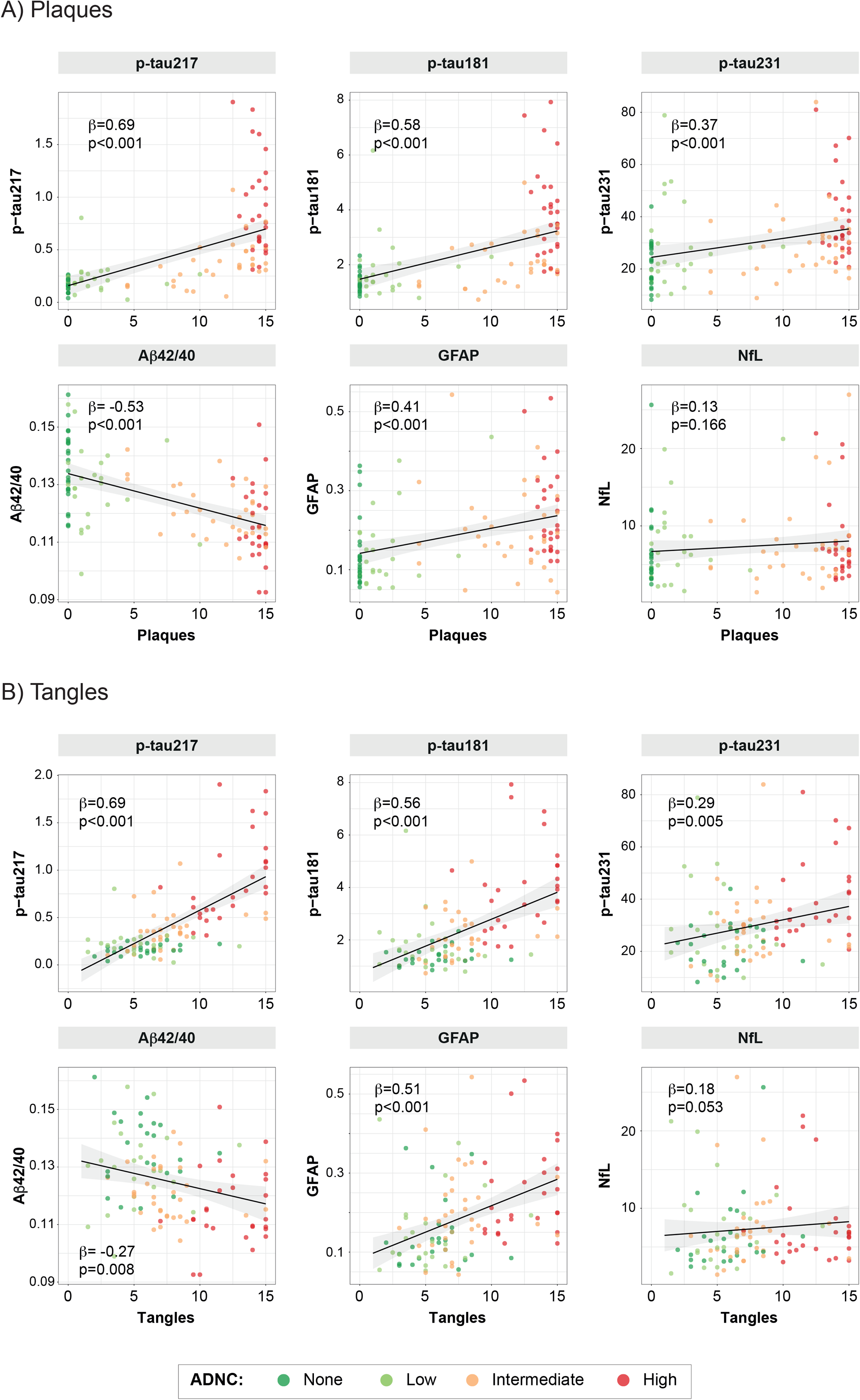
Associations between plasma biomarkers and amyloid plaque or neurofibrillary tau tangle loads. Black lines represent the association between plasma biomarkers and amyloid plaque (top) and tau tangle (bottom) loads after adjusting for covariates (age, sex, and time between blood sampling and death), but dots represent raw data. Datapoints are coloured based on the ADNC classification. Standardized betas and p-values of the association between plasma biomarkers and load of amyloid plaques or tau tangles are shown in the plot. Abbreviations: A β, amyloid-β; ADNC, Alzheimer’s disease neuropathologic change; GFAP, glial fibrillary acidic protein; NfL, neurofilament light; p-tau, phosphorylated tau.

Next, we investigated specific associations between plasma biomarkers and plaques and tangles including both measures of AD brain pathological changes as predictors in a combined model. In these models, all p-tau markers (p-tau217: β=0.46, p<0.001; p-tau181: β=0.41, p<0.001; p-tau231: β=0.32, p=0.007) and the Aβ42/40 ratio (β=-0.59, p<0.001) were significantly associated with plaques (Table 2). Plasma GFAP showed an association at statistical trend level (β=0.19, p=0.058). In this analysis, the Aβ42/40 ratio correlation coefficient with plaques was the highest, being significantly higher than that of p-tau231 (β_diff_= 0.27, p=0.022) but not than that of p-tau217 (β_diff_= 0.13, p=0.163). In addition, we observed that only p-tau217 (β=0.40, p<0.001), p-tau181 (β=0.30, p=0.004) and GFAP (β=0.39, p<0.001) were associated with tangles. The correlation coefficient of p-tau217 with tangles showed a trend of being significantly higher than that of p-tau181 (β_diff_= 0.11, p=0.052), when adjusting for the total amount of plaques.

We also calculated the partial R^2^ of load of amyloid plaques and load of tau tangles to each biomarker level. We observed that the highest partial R^2^ of amyloid was from the Aβ42/40 ratio and p-tau217 (partial R^2^=0.24 for both) although this represented a much higher percentage of the total R^2^ in the case of the Aβ42/40 ratio (77.6% *vs*. 40.4%, Figure 2 and Supplementary Table 1). The highest partial R^2^ for tau was observed in p-tau217 (partial R^2^=0.18, percentual partial R^2^=30.7%) but the percentual partial R^2^ was similar than that of plasma GFAP (partial R^2^=0.13, percentual R^2^=30.4%, Figure 2 and Supplementary Table 1).

**Figure 2.**
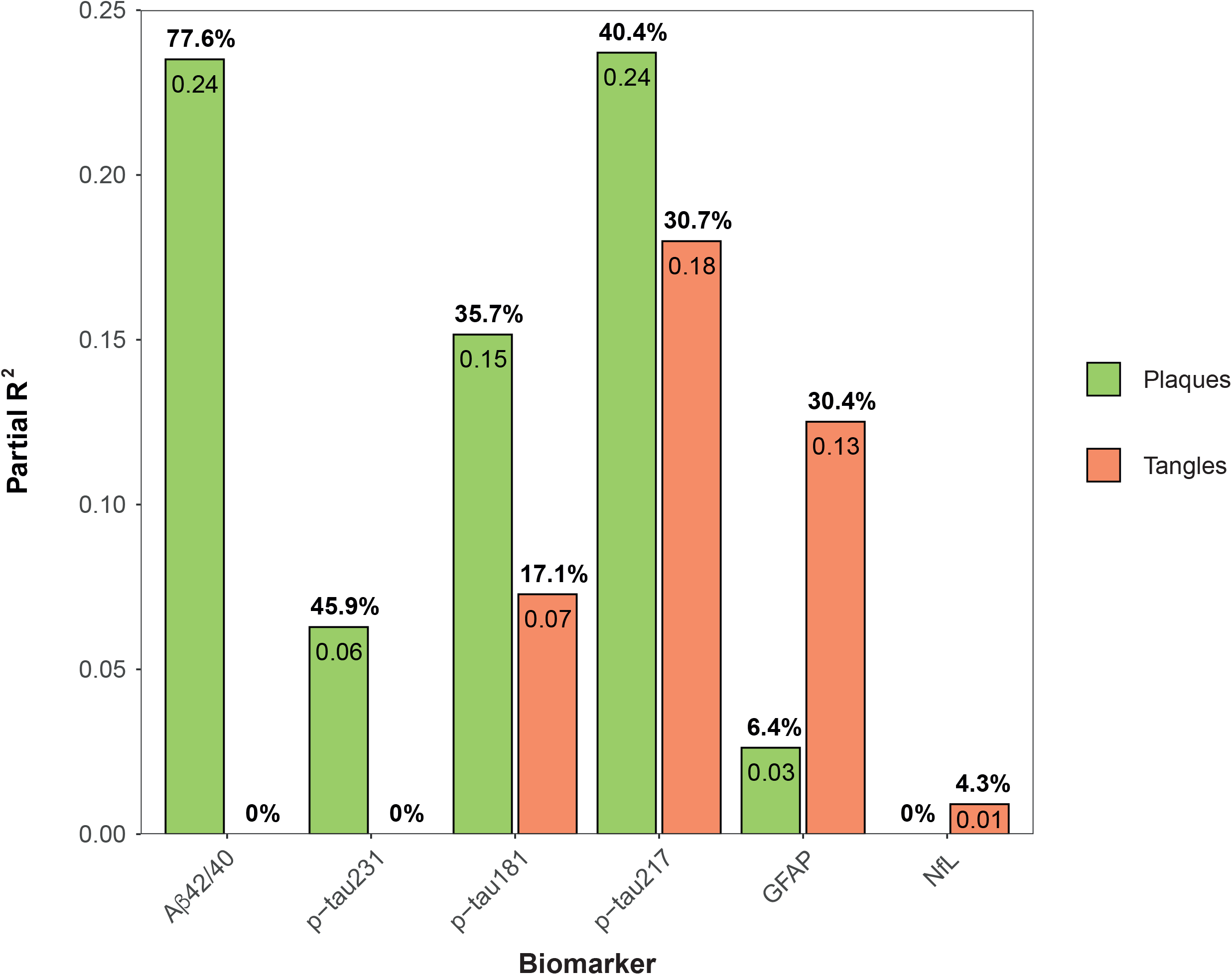
Contribution of amyloid plaque load and tau tangle load on plasma levels. Bars represent the partial R^2^ of amyloid plaque load (green) and tau tangle load (orange) on plasma levels. Linear regression models were used for these analyses with each biomarker as an outcome, in independent models, and both amyloid load and tau load as independent variables in a multivariable model. Age, sex, and time between blood sampling and death were included as covariates. Numbers inside the bars represent partial R^2^ and numbers above the bars represent the percentual partial R^2^ over the total R^2^ of each model (%partial R^2^ = 100*partial R^2^/total R^2^). Abbreviations: A β, amyloid-β; GFAP, glial fibrillary acidic protein; NfL, neurofilament light; p-tau, phosphorylated tau.

Finally, we investigated which combination of biomarkers better predicted plaques and tangles, independently. We found that the parsimonious model that better predicted load of amyloid plaques included both p-tau217 and the Aβ42/40 ratio (R^2^=0.57, Table 3), which was significantly better than the one only including p-tau217 based on AICc (ΔAICc=15.5). On the other hand, p-tau217 alone in the model was selected as the parsimonious model to predict load of neurofibrillary tangles (R^2^=0.50, Table 3).

**Table 3.** Parsimonious models to predict AD-related pathology. Parsimonious models were selected as those that better explained each AD-pathology measure with the smaller number of predictors based on the AICc criterion. Initial models included basic covariates (age, sex, and time between blood sampling) and all biomarkers that showed a significant association in the univariate analyses. Covariates were kept in the models even when they did not contribute to the model for a fair comparison to univariate analyses. Men are the reference sex group. Abbreviations: A β, amyloid-β; ADNC, Alzheimer’s disease neuropathologic change; AICc, corrected Akaike criterion, AUC, area under the curve; CI, confidence intervals; p-tau, phosphorylated tau.

| | $\beta$ [95%CI] | p-value<br>association | $R^2$ | AICc | AUC[95%CI] |
| --- | --- | --- | --- | --- | --- |
| Plaques |  |  |  |  |  |
| p-tau217 | 0.58 [0.44, 0.72] | <0.001 | 0.57 | 218.36 | - |
| Aβ42/40 | -0.32 [-0.46, -0.18] | <0.001 | | | |
| Age | 0.03 [-0.11, 0.16] | 0.705 |  |  |  |
| Sex | 0.01 [-0.26, 0.28] | 0.940 |  |  |  |
| Time blood-death | 0.00 [-0.14, 0.13] | 0.968 |  |  |  |
| Tangles |  |  |  |  |  |
| p-tau217 | 0.64 [0.50, 0.78] | <0.001 | 0.50 | 233.46 | - |
| Age | 0.02 [-0.12, 0.16] | 0.776 |  |  |  |
| Sex | -0.34 [-0.63, -0.05] | 0.020 |  |  |  |
| Time blood-death | 0.13 [-0.01, 0.27] | 0.077 |  |  |  |
| ADNC |  |  |  |  |  |
| p-tau217 | 1.83 [1.09, 2.79] | <0.001 | 0.66 | 93.77 | 0.90 [0.84, 0.96] |
| Aβ42/40 | -1.00 [-1.79, -0.29] | 0.008 | | | |
| Age | 0.29 [-0.26, 0.89] | 0.310 |  |  |  |
| Sex | 0.07 [-1.10, 1.26] | 0.908 |  |  |  |
| Time blood-death | 0.23 [-0.45, 0.96] | 0.513 |  |  |  |

### Prediction of neuropathological scales’ classification

First, we investigated differences in plasma levels by ADNC groups (as a four-level variable, *i*.*e*., none, low, intermediate, or high). All three p-tau plasma measures and the Aβ42/40 ratio showed significant differences between intermediate and high levels of ADNC. Plasma p-tau217 and the Aβ42/40 ratio levels were also significantly different between intermediate and low ADNC. However, we only found significant differences between none and low ADNC in plasma p-tau217 levels (Figure 3). Notably, p-tau217 also showed the highest fold-change among all ADNC consecutive levels (Supplementary Table 2).

**Figure 3.**
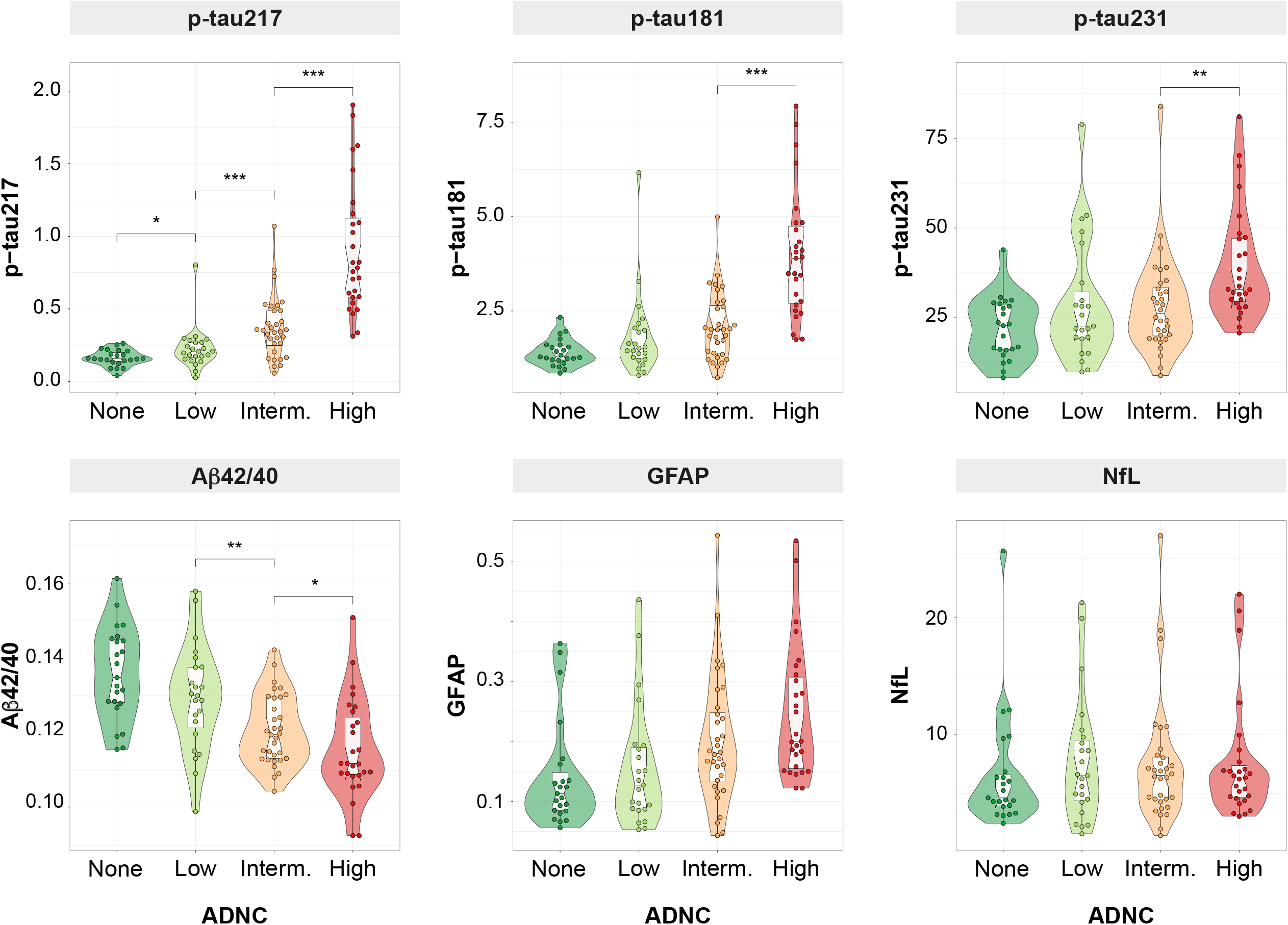
Plasma levels by ADNC classification. Groups were compared using pairwise Wilcoxon test as a *post hoc* comparison after testing tendency using a Kruskal-Wallis test. *Post hoc* comparisons were only performed between consecutive groups. *** p≤0.001; ** p≤0.010 ; * p≤0.050 Abbreviations: A β, amyloid-β; ADNC, Alzheimer’s disease neuropathologic change; GFAP, glial fibrillary acidic protein; NfL, neurofilament light; p-tau, phosphorylated tau.

Similarly, all biomarkers except NfL showed significant differences between sparse and moderate/frequent groups on CERAD’s classification. Only plasma p-tau217 showed differences between zero and sparse groups (Supplementary Figure 1). Regarding Braak staging, all biomarkers except NfL were significantly different when comparing 0-IV with V-VI groups (Supplementary Figure 2).

We next investigated the accuracy of each plasma biomarkers to predict presence of AD pathology as measured with the dichotomized ADNC (none/low *vs*. intermediate/high) classification. All biomarkers except NfL (AUC[95%CI]= 0.61 [0.50, 0.71], p=0.698) were predictive of presence of ADNC in the univariable analysis (0.88 ≥ AUC ≥ 0.72, Supplementary Table 3 and Figure 4). Plasma p-tau217 had the highest AUC (AUC[95%CI]= 0.88 [0.81-0.95]) of all biomarkers which was significantly higher than all others except for the Aβ42/40 ratio (AUC[95%CI]= 0.80 [0.72-0.89], p=0.099). We also repeated this analysis with CERAD (low/sparse *vs*. moderate/frequent) and Braak staging (0-IV *vs*. V-VI) classification. For CERAD, p-tau217 was also the best individual biomarker as per classification accuracy (AUC[95%CI]= 0.89 [0.83 – 0.96], p<0.001), comparable only to that of the Aβ42/40 ratio (AUC[95%CI]= 0.82 [0.74 – 0.90], p<0.001, Figure 4 and Supplementary Table 4). For Braak staging, p-tau217 again showed the highest accuracy (AUC[95%CI]= 0.93 [0.87 – 0.98], p<0.001) comparable only to that of GFAP (AUC[95%CI]= 0.86[0.79 – 0.94], p<0.001, Figure 4 and Supplementary Table 5).

**Figure 4.**
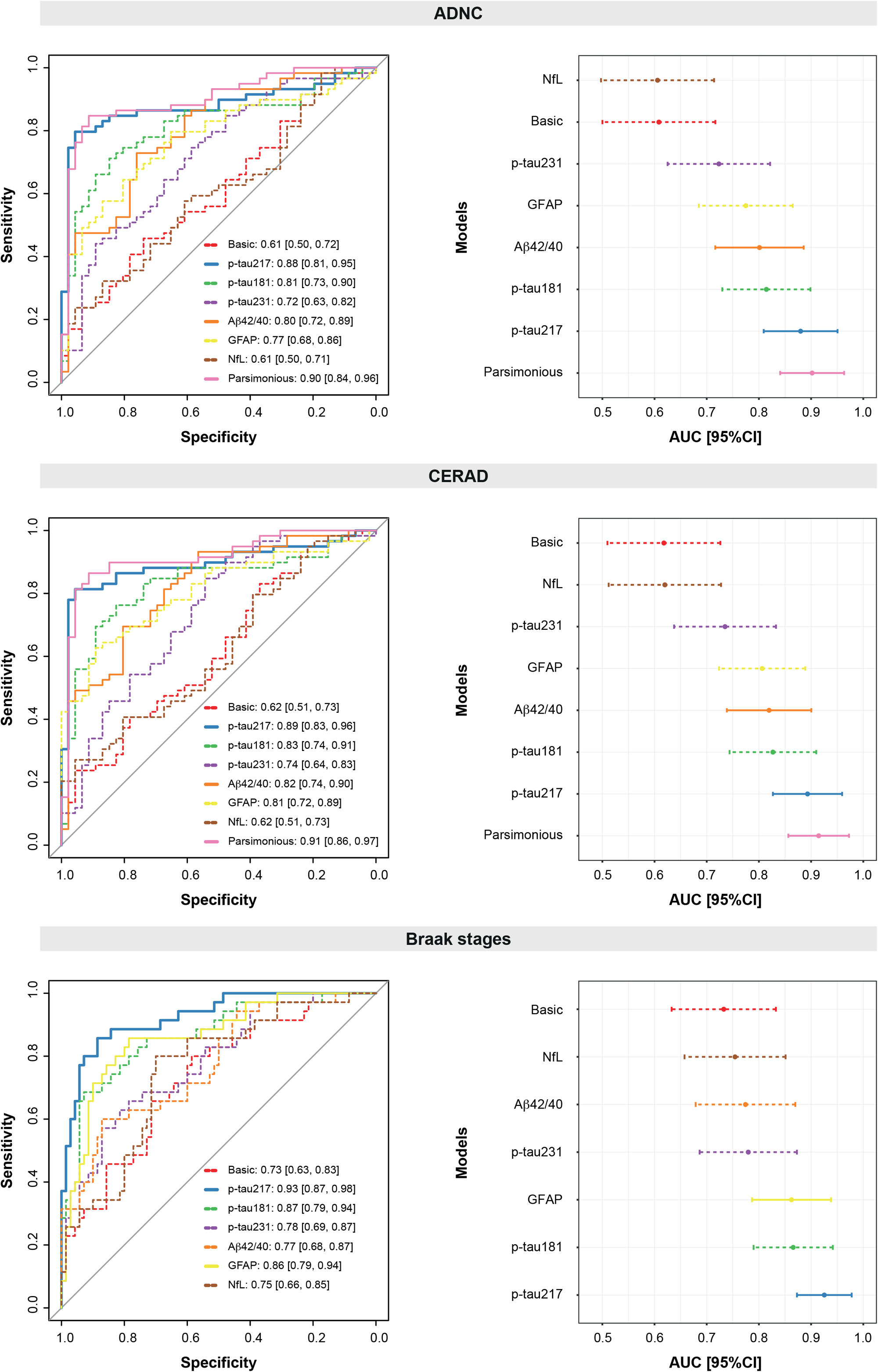
Plasma biomarkers for predicting neuropathological scales’ classification. ROC curves for all individual plasma biomarkers are shown in the left column and the correspondent AUC and 95%CI are shown in the right column. Models for all individual plasma biomarkers as well as the parsimonious (when available) model are shown. All models included: age, sex and time between blood sampling and death as covariates. The parsimonious model for ADNC and CERAD included p-tau217 and Aβ42/40 as predictors. The basic model includes only covariates. ADNC was dichotomized as negative (none/low) or positive (intermediate/high). CERAD was dichotomized as negative (low/sparse) or positive (moderate/frequent). Braak stages were dichotomized as negative (0-IV) or positive (V-VI). The individual biomarker with best performance is shown as a solid bold line. Dashed lines represent individual biomarkers with significant (p<0.05) lower AUC than the best individual biomarker (p-tau217 in all cases). Other models with solid lines represent AUC equivalent to that of the best individual biomarker. Abbreviations: A β, amyloid-β; ADNC, Alzheimer’s disease neuropathologic change; AUC, area under the curve, CERAD, Consortium to Establish a Registry for Alzheimer’s Disease; CI, confidence interval; GFAP, glial fibrillary acidic protein; NfL, neurofilament light; p-tau, phosphorylated tau; ROC, receiver operating characteristic.

Next, we determined the best combination of plasma biomarkers to predict presence of AD pathology. Both plasma p-tau217 and the Aβ42/40 ratio were included in the parsimonious model to predict presence of ADNC (AUC[95%CI]=0.90 [0.84, 0.96], Table 3 and Figure 4), but the AUC was not significantly higher than that of p-tau217 alone (p=0.124). With CERAD we observed a similar behaviour, with plasma p-tau217 and the Aβ42/40 ratio included in the parsimonious model (AUC[95%CI]=0.91 [0.86 – 0.97], Supplementary Table 6), although the AUC was not significantly better than that of p-tau217 alone (p=0.173). In the case of Braak staging, the parsimonious model only included p-tau217.

### Prediction of presence of co-pathologies

In this analysis, we investigated whether any of the available plasma biomarkers improved the basic models’ accuracy (only covariates) on predicting presence of co-pathologies, after adjusting for presence of intermediate/high ADNC. We observed that only plasma NfL significantly improved the prediction of presence of CWMR (AUC[95%CI]= 0.76 [0.66, 0.85]) compared to the basic model (AUC[95%CI]= 0.65 [0.54, 0.76], p=0.028, Supplementary Table 7 and Supplementary Figure 3A). In particular, participants with CWMR had significantly higher plasma NfL levels than those without (β=0.88, p=0.002, Figure 3B). No other biomarkers improved the prediction of presence of this nor any other co-pathology (Supplementary Tables 8-11 and Supplementary Figure 4). Raw distribution of plasma levels by presence of each co-pathology can be observed in Supplementary Figures 5-9.

### Use of the p-tau217/Aβ42 ratio

Given that the CSF p-tau/Aβ42 ratio is commonly used both in research and in clinical practice, we wanted to investigate whether a plasma p-tau/Aβ42 ratio would also be useful for predicting AD pathology. For this, we selected p-tau217 as it showed the highest associations in the previous analyses. First, we compared the accuracy of predicting plaques and tangles, independently, comparing parsimonious models including the plasma p-tau217/Aβ42 ratio as a possible independent variable to those obtained in the previous sections including p-tau217. We observed that the p-tau217/Aβ42 ratio was preferentially selected over p-tau217 in the models predicting plaques and tangles. Based on the AICc, we observed that models including the p-tau217/Aβ42 ratio were slightly, but significantly, better than those previously presented (plaques: R^2^ =0.60, AICc=210.1 *vs*. R^2^ =0.57, AICc=218.4; tangles: R^2^_p-tau217/Aβ42 ratio_=0.52, AICc=228.7 *vs*. R^2^_p-tau217_=0.50, AICc=233.5; Supplementary Table 12).

### Longitudinal associations between p-tau217 and p-tau181 with AD pathology

Finally, we investigated whether longitudinal changes in plasma p-tau217 and p-tau181 were associated with presence of AD pathology at death (median[range] timepoints: 2[2-5], mean(SD) time difference from first timepoint to death: 1378(1357) days). Details of these participants can be found in Supplementary Table 13. First, we observed that longitudinal increments of p-tau217 but not p-tau181 were associated with plaque burden (p-tau217: β=0.09, p=0.005; p-tau181: β=0.05, p=0.350, Table 4). In independent models, we observed that p-tau217 increments, but not those in p-tau181, were also associated with tangle load (p-tau217: β=0.09, p=0.004; p-tau181: β=0.08, p=0.094, Table 4).

**Table 4.** Associations between longitudinal changes of plasma biomarkers and presence of AD-related pathology at death. Linear mixed effect models were used to investigate these associations in independent models including: age at baseline and sex as covariates using and random intercepts and fixed time-slopes. The interaction between time and amyloid plaques, tau tangles or presence of ADNC were used as predictors, in independent models for both p-tau217 and p-tau181. Significant associations (p<0.05) between plasma biomarkers and presence of AD-related pathology are shown in bold. Abbreviations: ADNC, Alzheimer’s disease neuropathologic change; AICc, corrected Akaike criterion, CI, confidence intervals; p-tau, phosphorylated tau.

| Predictor | p-tau217 |  |  |  | p-tau181 |  |  |  |
| --- | --- | --- | --- | --- | --- | --- | --- | --- |
| | $\beta$ [95% CI] | p | R<sup>2</sup> | AICc | $\beta$ [95% CI] | p | R<sup>2</sup> | AICc |
| Plaques*time | 0.09<br>[0.03, 0.15] | **0.005** | 0.37 | 189.35 | 0.05<br>[-0.05, 0.15] | 0.350 | 0.30 | 255.72 |
| Tangles*time | 0.09<br>[0.03, 0.14] | **0.004** | 0.53 | 173.95 | 0.08<br>[-0.01, 0.17] | 0.094 | 0.35 | 248.79 |
| ADNC*time | 0.13<br>[0.02, 0.24] | **0.018** | 0.24 | 197.95 | 0.12<br>[-0.05, 0.29] | 0.152 | 0.21 | 258.79 |

Finally, we also investigated whether participants with intermediate/high ADNC pathology at death showed higher increments in p-tau levels compared with those with none/low ADNC pathology. We observed that participants with intermediate/high ADNC had significantly higher p-tau217, but not p-tau181, longitudinal increases (p-tau217: β=0.13, p=0.018; p-tau181: β=0.12, p=0.152, Table 4 and Supplementary Figure 10). These differences were observable up to seven years before death, as defined by non-overlapping 95%CIs.

## Discussion

In this study, we have investigated the specific associations between multiple plasma biomarkers, using high performing assays, and autopsy-assessed measures of AD pathology in a single cohort. Our main result was that the plasma Aβ42/40 ratio and p-tau231 were selectively associated with plaques, plasma GFAP only with tangles, whereas p-tau181 and, most strongly, p-tau217 were independently associated with both plaques and tangles. We also observed that the combination of p-tau217 and the Aβ42/40 ratio showed the highest accuracy to predict amyloid plaque load. On the other hand, p-tau217 alone was sufficient to accurately predict tau tangle load. Regarding co-pathologies, only the use of plasma NfL showed an improvement on predicting the presence of cerebral white matter rarefaction (CWMR), but no other biomarkers further improved this prediction nor any of any other co-pathology. Notably, the use of the plasma p-tau217/Aβ42 ratio showed slight, although significant, improvements compared to p-tau217 alone when assessing semi-quantitative measures of AD pathology. Finally, we observed that longitudinal increases of p-tau217, but not those of p-tau181, were significantly associated with presence of AD pathology at death, especially with tangle burden. Taken altogether, this study supports the use of plasma p-tau217 as the best biomarker for measuring AD-related pathology, supported by its independent associations with neuropathological measures of both plaques and tangles. Notably, the Aβ42/40 ratio may add independent information to that provided by p-tau217 for assessing amyloid pathology.

The main result of this study was the observation that plasma p-tau217 and plasma p-tau181 were specific markers of both amyloid plaques and tau tangles. A previous study with a subsample of the individuals included here (n=88) already suggested an independent association between plasma p-tau217 and the two main AD-related pathologies ^19^. The novelties of our study were to demonstrate that this dual association only occurred in p-tau217 and p-tau181. Further, we also observed that p-tau217 changed earlier along the ADNC scale (Figure 3), and also that longitudinal changes in plasma p-tau217, but not those of p-tau181, were associated with AD-related pathology. Altogether, our data suggest that plasma p-tau217 is the best suited plasma biomarker among the ones studied here to assess presence of AD-related pathology across the whole *continuum*. Although p-tau181 has shown very good performance as an AD biomarker,^4,8,9,16,39^ multiple (plasma and CSF) studies support our results that p-tau217 may be more useful than p-tau181, as it has shown better correlations with amyloid and tau pathology proxies, earlier change, and better diagnostic accuracy.^5,6,16,40–43^ Further, our longitudinal results suggest that the utilization of plasma p-tau217 in clinical trials may be useful not only as a pre-screening method, but also for disease monitoring, especially for those drugs targeting tau pathology.

However, while plasma p-tau217 and p-tau181 were associated with both plaques and tangles, the other studied biomarkers showed more specific associations. For instance, while plasma p-tau231 showed a significant correlation with tangle counts, it was also only associated with amyloid plaques in the multivariable analysis. Previous studies have suggested that p-tau231, both as a CSF and a plasma biomarker, may be an early AD marker tightly associated with amyloid pathology.^13,17,44–46^ Contrary to previous studies, we observed significantly lower associations with amyloid than that of p-tau217 and the Aβ42/40 ratio, and some elevated levels in subjects without neuropathological evidence of amyloid plaques (Figure 1). One possible explanation for the early increases observed here and in previous studies may be that they are related with soluble amyloid, which cannot be detected in our study and is presumably an earlier event in the Alzheimer’s *continuum*. We acknowledge that further research is needed to understand the relationship between this biomarker and actual pathology. Similarly, plasma Aβ42/40 ratio was also only associated with plaques in the multivariable analysis.^21,47^ However, the most important finding regarding the plasma Aβ42/40 ratio was that combining it with plasma p-tau217 could improve amyloid plaque assessment, replicating a previous result from our group when assessing amyloid positivity by CSF.^48^ Thus, our results support the use of the combination of plasma Aβ42/40 ratio and p-tau217 in clinical trials targeting amyloid pathology as a pre-screening method.

One surprising finding of our study was the specific association between plasma GFAP and tau tangles. Contrarily, previous studies have shown significant associations between this marker and amyloid pathology (as measured by CSF or PET), which were stronger than those with tau pathology (also measured with CSF or PET).^10,49^ Apart from the fact that GFAP levels were high in some cases with no or low amounts of plaques, which affects the correlations, two main points must be accounted when comparing these to our results. First, none of the aforementioned studies adjusted for tau pathology when assessing associations with amyloid. Following this approach, we also observed an association between plasma GFAP and plaques (univariable analysis). And second, that tau PET is known to not be sensitive to early tau pathology, which may have decreased the power to detect these associations in previous studies.^50–52^ On the other hand, recent studies have shown the association between higher levels of plasma GFAP and increased risk of clinical progression and steeper rates of cognitive decline (even after adjusting for amyloid),^53–55^ which supports a link with tau pathology given the known strong association between tau and clinical symptoms. Thus, our hypothesis is that plasma GFAP may be associated with AD pathology, but it may primarily relate to early deposited tau tangles.

The CSF p-tau/Aβ42 ratio has received a lot of attention in the recent years, both as a research and a clinical tool.^56–61^ Thus, we wanted to investigate whether a similar ratio would be also useful using plasma biomarkers. We observed that p-tau217/Aβ42 ratio slightly, but significantly, improved prediction accuracy to detect AD-related pathology. Although further investigation is needed, we suggest that this ratio may be useful to track AD pathology across the *continuum* due to its relationship with both main pathological hallmarks of AD, as well as better statistical characteristics of ratios, which can account for production/clearance participants’ inter-variability.^62,63^

Finally, we also investigated whether the levels of these biomarkers could be used to predict presence of common AD co-pathologies. Only plasma NfL significantly predicted the presence of CWMR (*i*.*e*., significantly improved the model with only covariates), with those subjects with presence of CWMR having higher plasma NfL levels. This is in agreement with NfL being a biomarker of non-specific axonal degeneration,^64,65^ but these results should be confirmed in an independent cohort. None of the biomarkers investigated could predict any of the other co-pathologies investigated (*i*.*e*., CAA, TDP-43, LBD and AGD), which replicates some of the results from a recent study in a different cohort with a subsample of the biomarkers described here.^17^ Given the results from our study and previous studies, we emphasize the urgent need of developing new biomarkers capable of measuring the presence of these and other common co-pathologies *in vivo* for a better diagnosis and prognosis for AD patients.

The main strength of this study was the availability of high performing assays of multiple plasma biomarkers, including the three main p-tau biomarkers phosphorylated at different sites, in a relatively large neuropathological cohort. Thus, we were able to directly compare specific associations between all of these biomarkers to gold standard measures of pathology in the same participants. Further, the use of semi-quantitative scores for measuring burden of AD pathology, compared with the typical dichotomous scales used, allowed us to perform more complex analyses. However, some limitations must be acknowledged. First, we recognize the small number of participants with intermediate levels of pathology and those with or without certain co-pathologies. Another limitation is the restricted number of participants in the longitudinal subsample, which may have reduced the power to find significant time interactions with plasma p-tau181. Regardless, our results point to stronger associations with p-tau217, which is in accordance with the cross-sectional analyses.

In conclusion, our study demonstrated that plasma p-tau217 and plasma p-tau181 are specific markers of both amyloid plaques and tau tangles, whereas the Aβ42/40 ratio and p-tau231 levels are markers strictly associated with plaques and GFAP with tangles. The combination of plasma p-tau217 and the Aβ42/40 ratio gives the highest accuracy for predicting amyloid plaque load, while p-tau217 alone may be sufficient to predict load of tangles. These results may be useful to design pre-screening strategies for clinical trials targeting amyloid and tau pathologies.

## Data Availability

All data produced in the present study are available upon reasonable request to the authors

## Acknowledgements

We want to thank all participants of the Arizona Study of Aging and Neurodegenerative Disorders and Brain and Body Donation Program (BBDP) at Banner Sun Health Research Institute and their families for their participation in the study. The Elecsys β-Amyloid(1–42), β-Amyloid(1–40), GFAP, and NfL prototype plasma immunoassays are intended for investigational purposes and are not currently approved for clinical use or commercially available. COBAS, COBAS E, and ELECSYS are trademarks of Roche.

