## Supplementary material for "Specific associations between plasma biomarkers and post-mortem amyloid plaque and neurofibrillary tau tangle loads"

**Supplementary Tables**

|  | Partial R^2^ plaques | % partial R^2^ plaques | | | Partial R^2^ tangles | | % partial R^2^ tangles |
| --- | --- | --- | --- | --- | --- | --- | --- |
| p-tau217 | 0.24 | | 40.4 | 0.18 | | 30.70 | |
| p-tau181 | 0.15 | | 35.7 | 0.07 | | 17.10 | |
| p-tau231 | 0.06 | | 45.9 | 0.00 | | 0.00 | |
| Aβ42/40 | 0.24 | | 77.6 | 0.00 | | 0.00 | |
| GFAP | 0.03 | | 6.4 | 0.13 | | 30.40 | |
| NfL | -0.01 | | 0.0 | 0.01 | | 4.30 | |

**Supplementary Table 1** **Contribution of amyloid plaque load and tau tangle load on plasma levels**

Linear regression models were used for these analyses with each biomarker as an outcome, in independent models, and both amyloid load and tau load as independent variables in a multivariable model. Age, sex, and time between blood sampling and death were included as covariates. Percentual partial R^2^ is calculated as the ratio of partial R^2^ over the total R^2^ of each model (%partial R^2^ = 100*partial R^2^/total R^2^).

Abbreviations: Aβ, amyloid-β; GFAP, glial fibrillary acidic protein; NfL, neurofilament light; p-tau, phosphorylated tau.

| ADNC | Fold increase none-low | p-value  none-low | Fold increase low-interm | p-value  low-interm | Fold increase interm-high | p-value interm-high |
| --- | --- | --- | --- | --- | --- | --- |
| p-tau217 | 0.39 | **0.025** | 0.61 | **<0.001** | 1.46 | **<0.001** |
| p-tau181 | 0.30 | 0.076 | 0.15 | **0.027** | 0.88 | **<0.001** |
| p-tau231 | 0.32 | 0.250 | -0.01 | 0.769 | 0.37 | **0.004** |
| Aβ42/40 | -0.05 | 0.070 | -0.06 | **0.014** | -0.04 | **0.040** |
| GFAP | 0.10 | 0.582 | 0.29 | 0.073 | 0.21 | 0.145 |
| NfL | 0.18 | 0.231 | -0.04 | 0.537 | 0.03 | 0.855 |

**Supplementary Table 2** **Plasma difference by ADNC levels**

Kruskal-Wallis tests were used to investigate differences in all plasma biomarkers by ADNC status. *Post hoc* analyses were performed with pairwise Wilcoxon rank sum tests among consecutive levels. Fold increases between consecutive levels were also calculated using the lowest level of each comparison as a reference. Significant differences (p<0.05) are shown in bold.

Abbreviations: Aβ amyloid-β; ADNC, Alzheimer’s disease neuropathologic change; GFAP, glial fibrillary acidic protein; NfL, neurofilament light; p-tau, phosphorylated tau.

| ADNC | β [95%CI] | p-value  β | AUC[95%CI] | R^2^ | AICc | p-value DeLong |
| --- | --- | --- | --- | --- | --- | --- |
| Basic | - | - | 0.61 [0.50, 0.72] | 0.07 | 146.60 | <0.001 |
| p-tau217 | 2.22 [1.43, 3.22] | **<0.001** | 0.88 [0.81, 0.95] | 0.63 | 99.37 | Ref |
| p-tau181 | 1.43 [0.84, 2.13] | **<0.001** | 0.81 [0.73, 0.9] | 0.42 | 119.96 | 0.002 |
| p-tau231 | 0.69 [0.24, 1.19] | **0.004** | 0.72 [0.63, 0.82] | 0.19 | 139.30 | 0.002 |
| Aβ42/40 | -1.24 [-1.86, -0.71] | **<0.001** | 0.80 [0.72, 0.89] | 0.35 | 123.71 | 0.123 |
| GFAP | 1.10 [0.56, 1.74] | **<0.001** | 0.77 [0.68, 0.86] | 0.29 | 130.78 | 0.015 |
| NfL | 0.09 [-0.36, 0.55] | 0.698 | 0.61 [0.50, 0.71] | 0.07 | 148.66 | <0.001 |

**Supplementary Table 3 Plasma biomarkers for predicting ADNC classification**

Generalized linear regression models were used to investigate these associations in independent models including: age, sex, and time between blood sampling and death as covariates. ADNC was used as dependent variable, dichotomized as negative (none/low) or positive (intermediate/high). The basic model includes only covariates. Significant associations (p<0.05) between plasma biomarkers and ADNC positivity are shown in bold. Differences between the AUCs were calculated using the DeLong test, with the highest AUC as reference (ref.), shown in the last column. Significant differences (p<0.05) can be understood as significantly weaker predictive power compared with that of p-tau217.

Abbreviations: Aβ, amyloid-β; AICc, corrected Akaike criterion, AUC, area under the curve; CERAD, Consortium to establish a registry for Alzheimer's disease; CI, confidence interval; GFAP, glial fibrillary acidic protein; NfL, neurofilament light; p-tau, phosphorylated tau.

| CERAD | β [95%CI] | p-value  β | AUC[95%CI] | R^2^ | AICc | p-value DeLong |
| --- | --- | --- | --- | --- | --- | --- |
| Basic | - | - | 0.62 [0.51, 0.73] | 0.08 | 145.98 | <0.001 |
| p-tau217 | 2.35 [1.53, 3.42] | **<0.001** | 0.89 [0.83, 0.96] | 0.66 | 96.12 | ref. |
| p-tau181 | 1.53 [0.92, 2.27] | **<0.001** | 0.83 [0.74, 0.91] | 0.45 | 116.69 | 0.003 |
| p-tau231 | 0.80 [0.34, 1.33] | **0.001** | 0.74 [0.64, 0.83] | 0.23 | 135.99 | 0.001 |
| Aβ42/40 | -1.37 [-2.04, -0.81] | **<0.001** | 0.82 [0.74, 0.90] | 0.40 | 119.36 | 0.104 |
| GFAP | 1.44 [0.82, 2.20] | **<0.001** | 0.81 [0.72, 0.89] | 0.40 | 122.14 | 0.039 |
| NfL | 0.26 [-0.19, 0.73] | 0.263 | 0.62 [0.51, 0.73] | 0.09 | 146.91 | <0.001 |

**Supplementary Table 4 Plasma biomarkers for predicting CERAD classification**

Generalized linear regression models were used to investigate these associations in independent models including: age, sex, and time between blood sampling and death as covariates. CERAD classification was used as dependent variable, dichotomized as negative (zero/sparse) or positive (moderate/frequent). The basic model includes only covariates. Significant associations (p<0.05) between plasma biomarkers and CERAD positivity are shown in bold. Differences between the AUCs were calculated using the DeLong test, with the highest AUC as reference (ref.), shown in the last column. Significant differences (p<0.05) can be understood as significantly weaker predictive power compared with that of p-tau217.

Abbreviations: Aβ, amyloid-β; AICc, corrected Akaike criterion; AUC, area under the curve; CERAD, Consortium to establish a registry for Alzheimer's disease; CI, confidence interval; GFAP, glial fibrillary acidic protein; NfL, neurofilament light; p-tau, phosphorylated tau.

| Braak staging | β [95%CI] | p-value  β | AUC[95%CI] | R^2^ | AICc | p-value DeLong |
| --- | --- | --- | --- | --- | --- | --- |
| Basic | - | - | 0.73 [0.63, 0.83] | 0.23 | 123.95 | <0.001 |
| p-tau217 | 2.44 [1.57, 3.57] | **<0.001** | 0.93 [0.87, 0.98] | 0.73 | 77.04 | ref. |
| p-tau181 | 1.46 [0.87, 2.17] | **<0.001** | 0.87 [0.79, 0.94] | 0.54 | 97.00 | 0.004 |
| p-tau231 | 0.65 [0.16, 1.20] | **0.013** | 0.78 [0.69, 0.87] | 0.33 | 119.31 | <0.001 |
| Aβ42/40 | -0.67 [-1.2, -0.20] | **0.008** | 0.77 [0.68, 0.87] | 0.32 | 118.31 | 0.001 |
| GFAP | 1.48 [0.83, 2.28] | **<0.001** | 0.86 [0.79, 0.94] | 0.51 | 102.07 | 0.061 |
| NfL | 0.48 [-0.03, 1.02] | 0.070 | 0.75 [0.66, 0.85] | 0.28 | 122.80 | <0.001 |

**Supplementary Table 5 Plasma biomarkers for predicting Braak staging classification**

Generalized linear regression models were used to investigate these associations in independent models including: age, sex and time between blood sampling and death as covariates. Braak staging was used as dependent variable, dichotomized as negative (0-IV) or positive (V-VI). The basic model includes only covariates. Significant associations (p<0.05) between plasma biomarkers and Braak staging positivity are shown in bold. Differences between the AUCs were calculated using the DeLong test, with the highest AUC as reference (ref.), shown in the last column. Significant differences (p<0.05) can be understood as significantly weaker predictive power compared with that of p-tau217.

Abbreviations: Aβ, amyloid-β; AICc, corrected Akaike criterion, AUC, area under the curve; CI, confidence interval; GFAP, glial fibrillary acidic protein; NfL, neurofilament light; p-tau, phosphorylated tau.

|  | β [95%CI] | p-value  association | R^2^ | AICc | AUC[95%CI] |
| --- | --- | --- | --- | --- | --- |
| CERAD | | | | | |
| p-tau217 | 1.94 [1.16, 2.93] | **<0.001** | 0.70 | 87.89 | 0.91 [0.86, 0.97] |
| Aβ42/40 | -1.22 [-2.08, -0.46] | **0.003** |  |  |  |
| Age | 0.26 [-0.32, 0.88] | 0.386 |  |  |  |
| Sex | -0.20 [-1.42, 1.02] | 0.750 |  |  |  |
| Time  blood-death | 0.25 [-0.47, 1.03] | 0.499 |  |  |  |
| Braak stages | | | | | |
| p-tau217 | 2.44 [1.57, 3.57] | **<0.001** | 0.73 | 77.04 | 0.93 [0.87, 0.98] |
| Age | -0.31 [-0.98, 0.33] | 0.347 |  |  |  |
| Sex | -1.78 [-3.34, -0.45] | **0.014** |  |  |  |
| Time  blood-death | 0.83 [0.10, 1.67] | **0.035** |  |  |  |

**Supplementary Table 6** **Parsimonious models to predict CERAD and Braak staging classification**

Parsimonious models were selected as those that better explained each AD-related scale with the smaller number of predictors based on the AICc criterion. Initial models included basic covariates (age, sex, and time between blood sampling) and all biomarkers that showed a significant association in the univariate analyses. Men are the reference sex group. CERAD classification was used as dependent variable, dichotomized as negative (zero/sparse) or positive (moderate/frequent). Braak staging was used as dependent variable, dichotomized as negative (0-IV) or positive (V-VI).

Abbreviations: Aβ, amyloid-β; AICc, corrected Akaike criterion, AUC, area under the curve; CERAD, Consortium to Establish a Registry for Alzheimer's Disease; CI, confidence intervals; p-tau, phosphorylated tau.

| CWMR | β [95%CI] | p-value  β | AUC[95%CI] | R^2^ | AICc | p-value DeLong |
| --- | --- | --- | --- | --- | --- | --- |
| Basic | - | - | 0.65 [0.54, 0.76] | 0.10 | 146.18 | Ref. |
| p-tau217 | 0.58 [0.04, 1.18] | 0.043 | 0.70 [0.60, 0.80] | 0.16 | 144.02 | 0.145 |
| p-tau181 | 0.28 [-0.21, 0.78] | 0.271 | 0.67 [0.57, 0.78] | 0.12 | 147.20 | 0.359 |
| p-tau231 | 0.01 [-0.42, 0.45] | 0.957 | 0.65 [0.55, 0.76] | 0.10 | 148.43 | 0.548 |
| Aβ42/40 | 0.07 [-0.40, 0.55] | 0.774 | 0.66 [0.55, 0.76] | 0.10 | 148.35 | 0.527 |
| GFAP | 0.40 [-0.08, 0.92] | 0.111 | 0.68 [0.58, 0.79] | 0.13 | 145.80 | 0.303 |
| NfL | 0.88 [0.36, 1.47] | 0.002 | 0.76 [0.66, 0.85] | 0.25 | 136.57 | **0.028** |

**Supplementary Table 7** **Plasma biomarkers for predicting presence of cerebral white matter rarefaction**

Generalized linear regression models were used to investigate these associations in independent models including: age, sex, time between blood sampling and death, and ADNC status, as a dichotomous variable, as covariates. CWMR was used as dependent variable, dichotomized as negative or positive. ADNC was dichotomized as negative (none/low) or positive (intermediate/high). The basic model includes only covariates. Differences between the AUCs were calculated using the DeLong test, with the AUC from the basic model as reference (ref.), shown in the last column. Significant differences (p<0.05) can be understood as significantly greater predictive power compared to that of only using covariates and are shown in bold.

Abbreviations: Aβ, amyloid-β; AICc, corrected Akaike criterion, AUC, area under the curve; CI, confidence interval; CWMR, cerebral white matter rarefaction; GFAP, glial fibrillary acidic protein; NfL, neurofilament light; p-tau, phosphorylated tau.

| CAA | β [95%CI] | p-value  β | AUC[95%CI] | R^2^ | AICc | p-value DeLong |
| --- | --- | --- | --- | --- | --- | --- |
| Basic | - | - | 0.82 [0.73, 0.91] | 0.36 | 113.41 | ref. |
| p-tau217 | 0.61 [-0.05, 1.32] | 0.078 | 0.85 [0.77, 0.93] | 0.40 | 112.38 | 0.209 |
| p-tau181 | 0.49 [-0.13, 1.15] | 0.129 | 0.84 [0.76, 0.92] | 0.39 | 113.25 | 0.297 |
| p-tau231 | -0.01 [-0.52, 0.49] | 0.970 | 0.82 [0.73, 0.91] | 0.36 | 115.66 | 0.660 |
| Aβ42/40 | -0.66 [-1.30, -0.08] | 0.032 | 0.84 [0.76, 0.92] | 0.42 | 110.60 | 0.225 |
| GFAP | 0.09 [-0.51, 0.69] | 0.764 | 0.82 [0.73, 0.91] | 0.36 | 115.57 | 0.860 |
| NfL | -0.18 [-0.76, 0.38] | 0.527 | 0.81 [0.72, 0.90] | 0.37 | 115.26 | 0.587 |

**Supplementary Table 8** **Plasma biomarkers for predicting presence of CAA**

Generalized linear regression models were used to investigate these associations in independent models including: age, sex, time between blood sampling and death, amyloid plaque and tau tangle measures as covariates. Presence or absence of CAA was used as dependent variable, dichotomized as negative (0-1) or positive (2-3). The basic model includes only covariates. Differences between the AUCs were calculated using the DeLong test, with the AUC from the basic model as reference (ref.), shown in the last column. Significant differences (p<0.05) can be understood as significantly greater predictive power compared with that of only using covariates.

Abbreviations: Aβ, amyloid-β; AICc, corrected Akaike criterion, CAA, cerebral amyloid angiopathy; AUC, area under the curve; CI, confidence interval; GFAP, glial fibrillary acidic protein; NfL, neurofilament light; p-tau, phosphorylated tau.

| LBD | β [95%CI] | p-value  β | AUC[95%CI] | R^2^ | AICc | p-value DeLong |
| --- | --- | --- | --- | --- | --- | --- |
| Basic | - | - | 0.65 [0.51, 0.8] | 0.08 | 111.53 | ref. |
| p-tau217 | 0.12 [-0.52, 0.81] | 0.727 | 0.64 [0.50, 0.79] | 0.08 | 113.65 | 0.228 |
| p-tau181 | 0.07 [-0.54, 0.66] | 0.829 | 0.65 [0.50, 0.79] | 0.08 | 113.73 | 0.495 |
| p-tau231 | 0.04 [-0.49, 0.56] | 0.890 | 0.65 [0.50, 0.79] | 0.08 | 113.76 | 0.246 |
| Aβ42/40 | 0.67 [0.07, 1.33] | 0.036 | 0.70 [0.58, 0.83] | 0.15 | 108.98 | 0.364 |
| GFAP | 0.02 [-0.57, 0.61] | 0.954 | 0.65 [0.51, 0.80] | 0.08 | 113.77 | 0.847 |
| NfL | 0.01 [-0.55, 0.56] | 0.965 | 0.65 [0.51, 0.80] | 0.08 | 113.78 | 0.339 |

**Supplementary Table 9** **Plasma biomarkers for predicting presence of LBD**

Generalized linear regression models were used to investigate these associations in independent models including: age, sex, time between blood sampling and death, amyloid plaque and tau tangle measures as covariates. Presence or absence of LBD was used as dependent variable, dichotomized as negative or positive. The basic model includes only covariates. Differences between the AUCs were calculated using the DeLong test, with the AUC from the basic model as reference (ref.), shown in the last column. Significant differences (p<0.05) can be understood as significantly greater predictive power compared with that of only using covariates.

Abbreviations: Aβ, amyloid-β; AICc, corrected Akaike criterion, AUC, area under the curve; CI, confidence interval; GFAP, glial fibrillary acidic protein; LBD, Lewy body disease; NfL, neurofilament light; p-tau, phosphorylated tau.

| TDP-43 | β [95%CI] | p-value  β | AUC[95%CI] | R^2^ | AICc | p-value DeLong |
| --- | --- | --- | --- | --- | --- | --- |
| Basic | - | - | 0.60 [0.43, 0.77] | 0.05 | 82.35 | - |
| p-tau217 | 0.59 [-0.14, 1.42] | 0.134 | 0.70 [0.56, 0.84] | 0.11 | 82.40 | 0.106 |
| p-tau181 | 0.35 [-0.30, 1.05] | 0.296 | 0.65 [0.50, 0.81] | 0.08 | 83.72 | 0.215 |
| p-tau231 | -0.01 [-0.57, 0.55] | 0.977 | 0.60 [0.44, 0.77] | 0.05 | 84.84 | 0.528 |
| Aβ42/40 | -0.32 [-1.00, 0.32] | 0.337 | 0.62 [0.46, 0.79] | 0.08 | 83.90 | 0.564 |
| GFAP | 0.03 [-0.62, 0.68] | 0.936 | 0.60 [0.43, 0.77] | 0.05 | 84.84 | 0.892 |
| NfL | 0.20 [-0.38, 0.78] | 0.498 | 0.63 [0.47, 0.78] | 0.06 | 84.39 | 0.498 |

**Supplementary Table 10** **Plasma biomarkers for predicting presence of TDP-43 pathology**

Generalized linear regression models were used to investigate these associations in independent models including: age, sex, time between blood sampling and death, amyloid plaque and tau tangle measures as covariates. Presence or absence of TDP-43 pathology was used as dependent variable, dichotomized as negative or positive. Differences between the AUCs were calculated using the DeLong test, with the AUC from the basic model as reference (ref.), shown in the last column. Significant differences (p<0.05) can be understood as significantly greater predictive power compared with that of only using covariates.

Abbreviations: Aβ, amyloid-β; AICc, corrected Akaike criterion, AUC, area under the curve; CI, confidence interval; GFAP, glial fibrillary acidic protein; NfL, neurofilament light; p-tau, phosphorylated tau; TDP-43, TAR DNA binding protein-43.

| AGD | β [95%CI] | p-value  β | AUC[95%CI] | R^2^ | AICc | p-value DeLong |
| --- | --- | --- | --- | --- | --- | --- |
| Basic | *-* | *-* | 0.53 [0.41, 0.66] | 0.01 | 131.31 | ref. |
| p-tau217 | -0.60 [-1.21, -0.04] | 0.042 | 0.65 [0.54, 0.77] | 0.07 | 129.22 | 0.073 |
| p-tau181 | -0.35 [-0.92, 0.18] | 0.204 | 0.60 [0.48, 0.72] | 0.03 | 131.89 | 0.257 |
| p-tau231 | -0.27 [-0.76, 0.20] | 0.274 | 0.58 [0.47, 0.70] | 0.03 | 132.35 | 0.370 |
| Aβ42/40 | 0.53 [0.00, 1.10] | 0.058 | 0.63 [0.52, 0.75] | 0.06 | 129.73 | 0.180 |
| GFAP | -0.11 [-0.66, 0.42] | 0.684 | 0.54 [0.41, 0.67] | 0.01 | 133.40 | 0.880 |
| NfL | -0.22 [-0.76, 0.28] | 0.406 | 0.55 [0.42, 0.67] | 0.02 | 132.86 | 0.775 |

**Supplementary Table 11** **Plasma biomarkers for predicting presence of AGD**

Generalized linear regression models were used to investigate these associations in independent models including: age, sex, time between blood sampling and death, amyloid plaque and tau tangle measures as covariates. Presence or absence of AGD pathology was used as dependent variable, dichotomized as negative or positive. The basic model includes only covariates. Differences between the AUCs were calculated using the DeLong test, with the AUC from the basic model as reference (ref.), shown in the last column. Significant differences (p<0.05) can be understood as significantly greater predictive power compared with that of only using covariates.

Abbreviations: Aβ, amyloid-β; AGD, argyrophilic grain disease; AICc, corrected Akaike criterion, AUC, area under the curve; CI, confidence interval; GFAP, glial fibrillary acidic protein; NfL, neurofilament light; p-tau, phosphorylated tau.

|  | β [95%CI] | p-value  association | R^2^ | AICc | AUC[95%CI] |
| --- | --- | --- | --- | --- | --- |
| Plaques | | | | | |
| p-tau217/Aβ42 ratio | 0.63 [0.49, 0.78] | **<0.001** | 0.60 | 210.1 | NA |
| Aβ42/40 | -0.26 [-0.4, -0.12] | **<0.001** |  |  |  |
| Age | 0.08 [-0.05, 0.21] | 0.202 |  |  |  |
| Sex | 0.01 [-0.25, 0.27] | 0.929 |  |  |  |
| Time  blood-death | -0.02 [-0.15, 0.11] | 0.804 |  |  |  |
| Tangles | | | | | |
| p-tau217/Aβ42 ratio | 0.66 [0.52, 0.80] | **<0.001** | 0.52 | 228.7 | NA |
| Age | 0.08 [-0.06, 0.22] | 0.281 |  |  |  |
| Sex | -0.33 [-0.61, -0.05] | **0.023** |  |  |  |
| Time  blood-death | 0.11 [-0.03, 0.26] | 0.113 |  |  |  |
| ADNC | | | | | |
| p-tau217/Aβ42 ratio | 2.65 [1.75, 3.82] | **<0.001** | 0.70 | 88.71 | 0.91 [0.85, 0.97] |
| Age | 0.56 [0, 1.19] | 0.061 |  |  |  |
| Sex | -0.29 [-1.47, 0.85] | 0.618 |  |  |  |
| Time  blood-death | 0.32 [-0.43, 1.1] | 0.413 |  |  |  |

**Supplementary Table 12** **Parsimonious models to predict AD-related pathology using the p-tau217/Aβ42 ratio**

Parsimonious models were selected as those that better explained each AD-pathology measure with the smaller number of predictors based on the AICc criterion. Initial models included basic covariates (age, sex, and time between blood sampling) and all biomarkers that showed a significant association in the univariate analyses including both p-tau217 alone and p-tau217/Aβ42 ratio. Men are the reference sex group.

Abbreviations: Aβ, amyloid-β; AICc, corrected Akaike criterion, ADNC, Alzheimer’s disease neuropathologic change; AUC, area under the curve; CI, confidence interval; p-tau, phosphorylated tau.

|  | Overall (n=48) | ADNC – negative (n=22) | ADNC – positive (n=26) |
| --- | --- | --- | --- |
| Age at baseline, mean(SD) | 85.6 (7.99) | 84.9 (6.92) | 86.2 (8.88) |
| Women, n(%) | 21 (43.8%) | 10 (45.5%) | 11 (42.3%) |
| APOE-e4 carrier, n (%) | 12 (25.0%) | 1 (4.5%) | 11 (42.3%) |
| Plaque total, mean(SD) | 6.47 (5.98) | 0.818 (1.22) | 11.3 (3.76) |
| CERAD moderate/frequent, n(%) | 26 (54.2%) | 0 (0%) | 26 (100%) |
| Tangle total, mean(SD) | 6.94 (2.69) | 5.86 (1.68) | 7.85 (3.07) |
| Braak stage, n(%) |  |  |  |
| I-II | 1 (2.1%) | 1 (4.5%) | 0 (0%) |
| III-IV | 40 (83.3%) | 20 (90.9%) | 20 (76.9%) |
| V-VI | 7 (14.6%) | 1 (4.5%) | 6 (23.1%) |
| Timepoints, median[range] | 2 [2-5] | 2 [2-5] | 2 [2-4] |
| Time difference, days, mean(SD) | 1,378 (1,357) | 1,411 (1,398) | 1,350 (1,349) |

**Supplementary Table 13** **Demographic characteristics of the longitudinal subsample**

ADNC was dichotomized as: negative (none/low) and positive (intermediate/high).

Abbreviations: ADNC, Alzheimer’s disease neuropathologic change; CERAD, Consortium to Establish a Registry for Alzheimer's Disease.

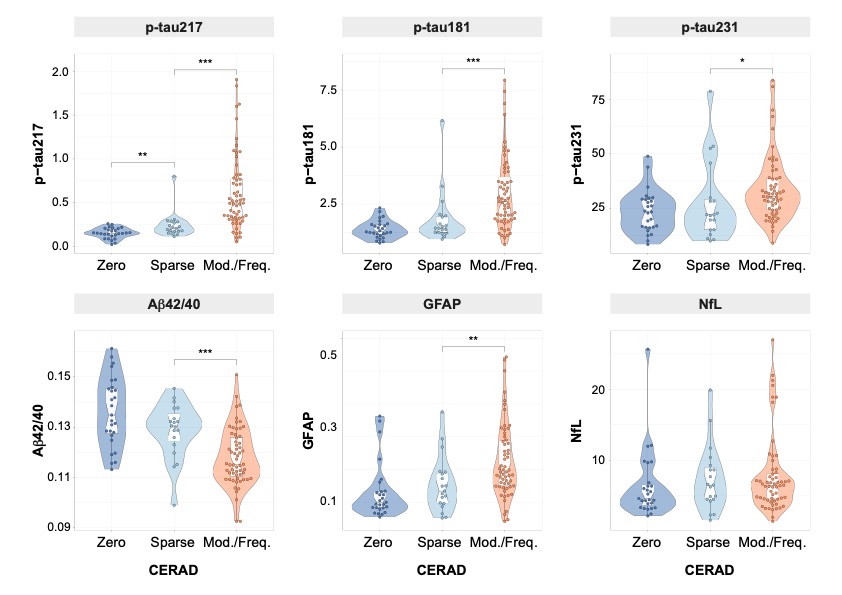
**Supplementary Figures**

**Supplementary Figure 1** **Plasma levels by CERAD classification**

Groups were compared using a Kruskal-Wallis test.

*** p<0.001; ** p<0.010 ; * p<0.050

Abbreviations: Aβ, amyloid-β; CAA, cerebral amyloid angiopathy; CERAD, Consortium to Establish a Registry for Alzheimer's Disease; GFAP, glial fibrillary acidic protein; NfL, neurofilament light; p-tau, phosphorylated tau.

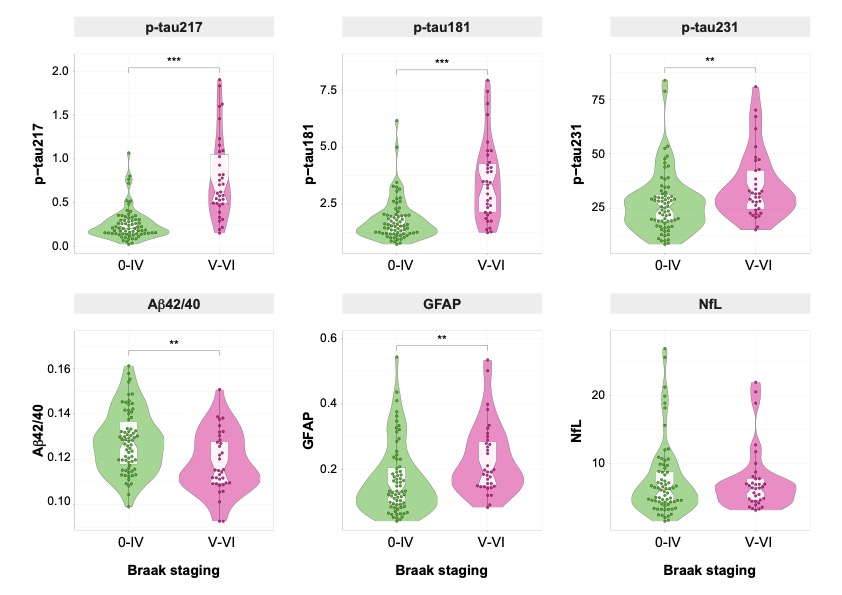

**Supplementary Figure 2** **Plasma levels by Braak staging classification**

Groups were compared using a Kruskal-Wallis test.

*** p<0.001; ** p<0.010 ; * p<0.050

Abbreviations: Aβ, amyloid-β; CAA, cerebral amyloid angiopathy; CERAD, Consortium to Establish a Registry for Alzheimer's Disease; GFAP, glial fibrillary acidic protein; NfL, neurofilament light; p-tau, phosphorylated tau.

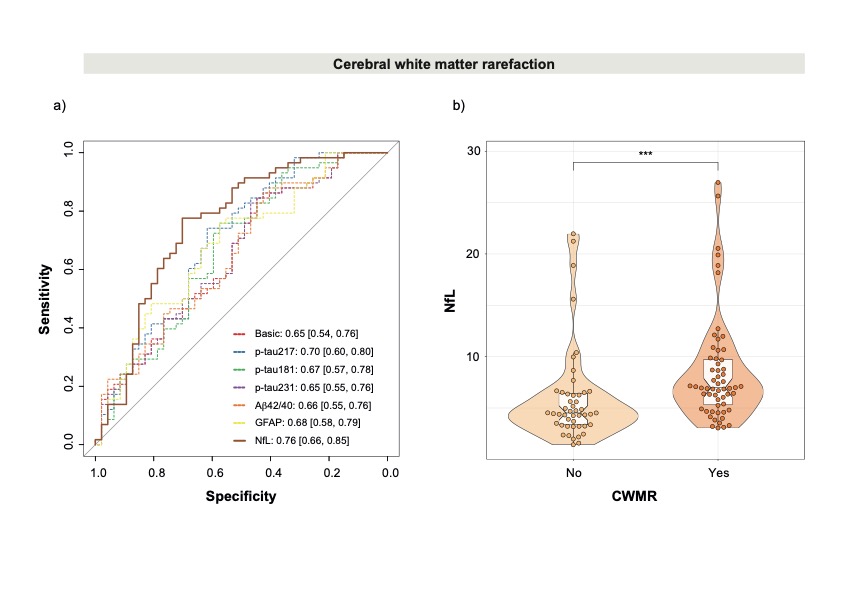

**Supplementary Figure 3** **Plasma biomarkers for predicting presence of cerebral white matter rarefaction**

ROC curves for all individual plasma biomarkers are shown in the left panel (a). In the ROC curves, all models included: age, sex, time between blood sampling and death, and presence of ADNC as a dichotomous variable as covariates. CWMR was used as dependent variable, dichotomized as negative or positive. ADNC was dichotomized as negative (none/low) or positive (intermediate/high). The basic model includes only covariates. AUCs and 95%CI are shown in the figure. The individual biomarker with best performance is shown as a solid bold line. Dashed lines represent individual biomarkers with significantly (p<0.05) lower AUC than the best individual biomarker. Only the addition of plasma NfL showed a significantly higher AUC than that of the basic model. Boxplot of plasma NfL levels by presence/absence of CWMR is shown in the right panel (b).

Abbreviations: Aβ, amyloid-β; ADNC, Alzheimer’s disease neuropathologic change; AUC, area under the curve; CI, confidence interval; CWMR, cerebral white matter rarefaction; GFAP, glial fibrillary acidic protein; NfL, neurofilament light; p-tau, phosphorylated tau; ROC, receiver operating characteristic.

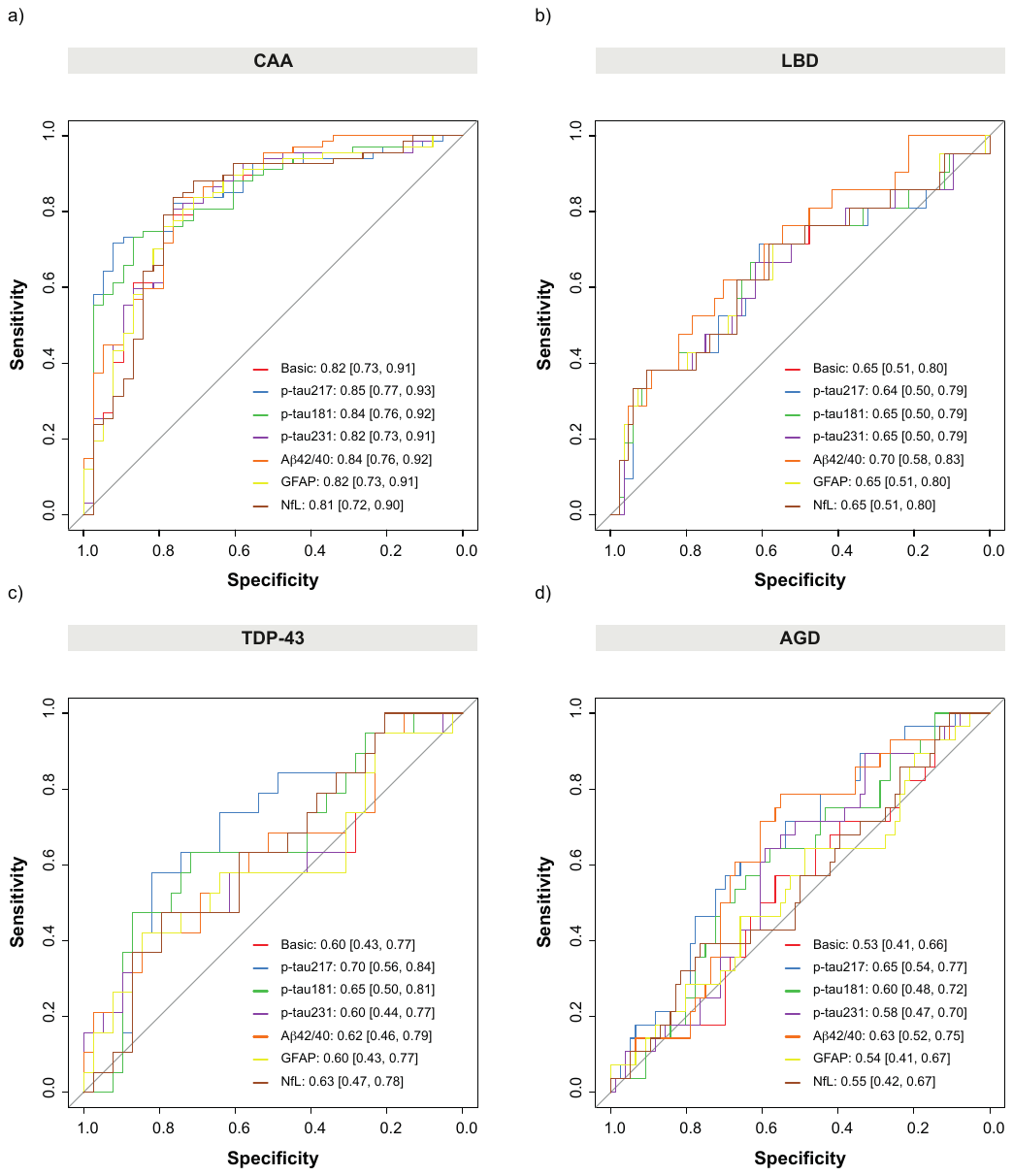

**Supplementary Figure 4** **ROC curves showing diagnostic accuracy of plasma biomarkers for predicting presence of co-pathologies**

ROC curves for predicting: CAA (a), LBD (b), TDP-43 (c) and AGD (d). All models included: age, sex, time between blood sampling, and death and presence of ADNC as a dichotomous variable as covariates. ADNC was dichotomized as negative (none/low) or positive (intermediate/high). The basic model includes only covariates. AUCs and 95%CI are shown in the figure.

Abbreviations: Aβ, amyloid-β; AUC, area under the curve, CAA, cerebral amyloid angiopathy; CI, confidence interval; GFAP, glial fibrillary acidic protein; LBD, Lewy body disease; NfL, neurofilament light; p-tau, phosphorylated tau; ROC, receiver operating characteristic; TDP-43, TAR DNA binding protein 43.

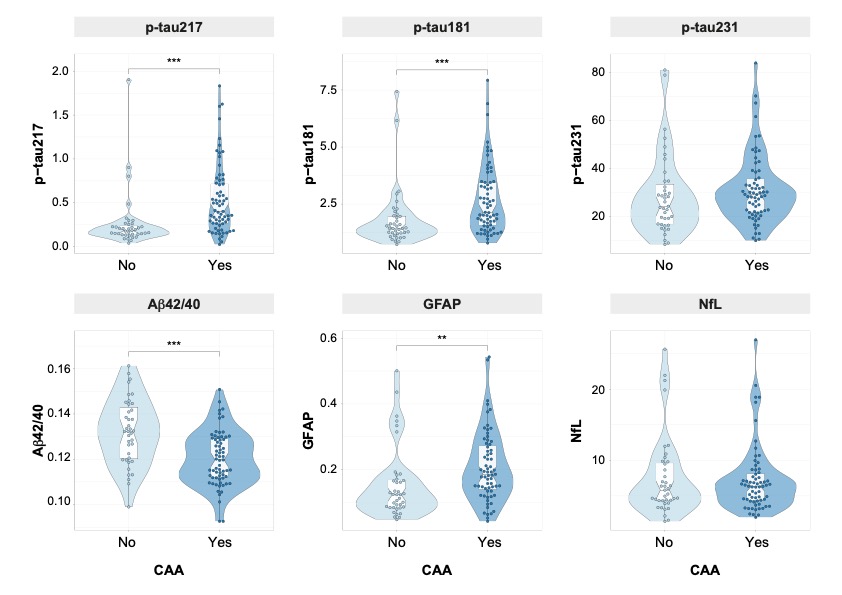

**Supplementary Figure 5** **Plasma levels by presence or absence of CAA**

Groups were compared using a Kruskal-Wallis test. None of these differences remained significant after adjusting for covariates (Supplementary Table 8).

*** p<0.001; ** p<0.010 ; * p<0.050

Abbreviations: Aβ, amyloid-β; CAA, cerebral amyloid angiopathy; GFAP, glial fibrillary acidic protein; NfL, neurofilament light; p-tau, phosphorylated tau.

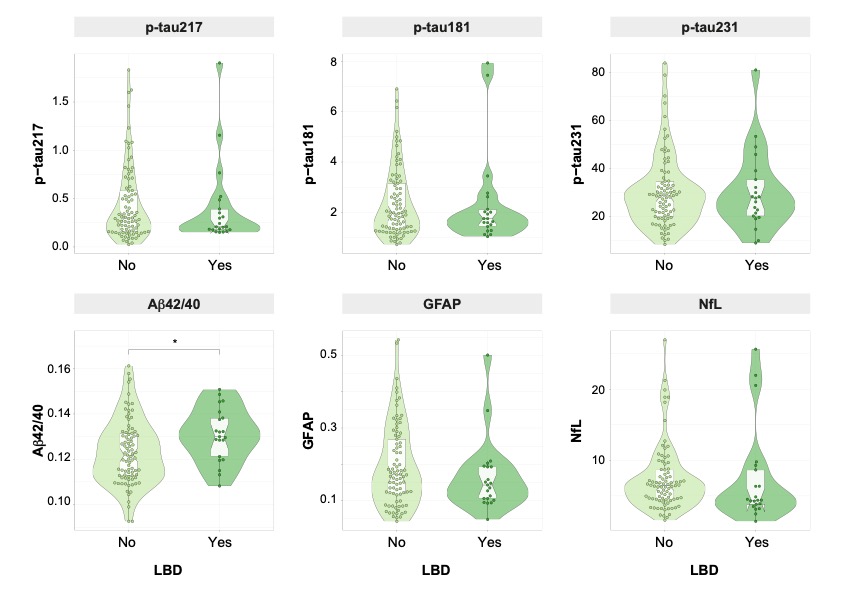

**Supplementary Figure 6 Plasma levels by presence or absence of LBD**

Groups were compared using a Kruskal-Wallis test. Only Aβ42/40 differences remained significant after adjusting for covariates (Supplementary Table 9).

*** p<0.001; ** p<0.010 ; * p<0.050

Abbreviations: Aβ, amyloid-β; GFAP, glial fibrillary acidic protein; LBD, Lewy body disease; NfL, neurofilament light; p-tau, phosphorylated tau.

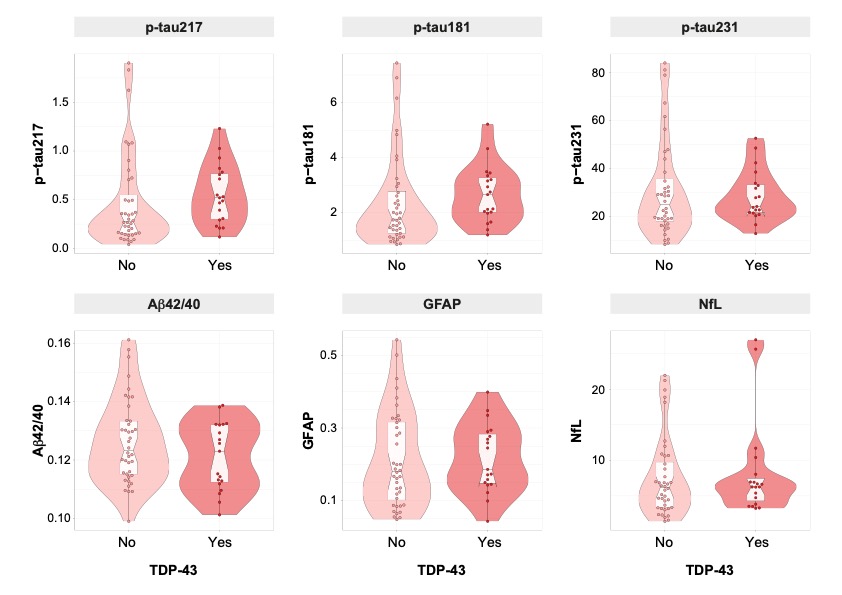

**Supplementary Figure 7** **Plasma levels by presence or absence of TDP-43 pathology**

Groups were compared using a Kruskal-Wallis test. None of these differences remained significant after adjusting covariates (Supplementary Table 10).

*** p<0.001; ** p<0.010 ; * p<0.050

Abbreviations: Aβ, amyloid-β; GFAP, glial fibrillary acidic protein; LBD, Lewy body disease; NfL, neurofilament light; p-tau, phosphorylated tau.

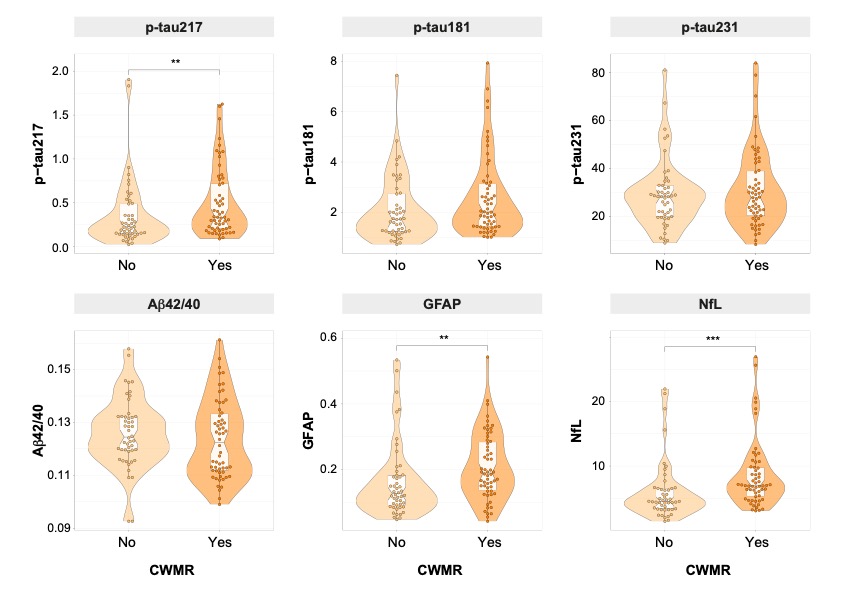

**Supplementary Figure 8** **Plasma levels by presence or absence of CWMR**

Groups were compared using a Kruskal-Wallis test. Differences of plasma p-tau217 and NfL levels remained significant after adjusting for covariates (Supplementary Table 7).

*** p<0.001; ** p<0.010 ; * p<0.050

Abbreviations: Aβ, amyloid-β; CWMR, cerebral white matter rarefaction; GFAP, glial fibrillary acidic protein; NfL, neurofilament light; p-tau, phosphorylated tau.

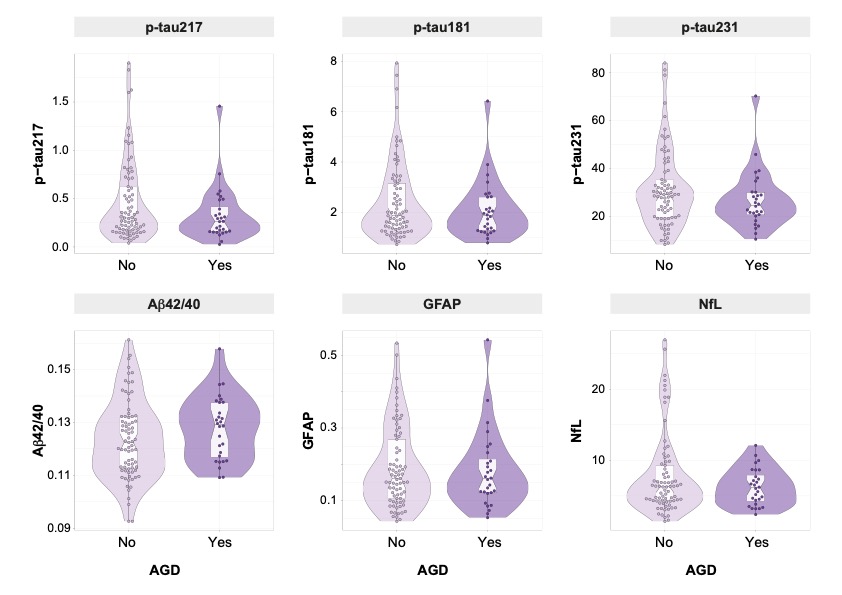

**Supplementary Figure 9** **Plasma levels by presence or absence of AGD**

Groups were compared using a Kruskal-Wallis test. Differences of plasma p-tau217 levels became significant after adjusting for covariates (Supplementary Table 11).

*** p<0.001; ** p<0.010 ; * p<0.050

Abbreviations: Aβ, amyloid-β; AGD, argyrophilic grain disease; GFAP, glial fibrillary acidic protein; NfL, neurofilament light; p-tau, phosphorylated tau.

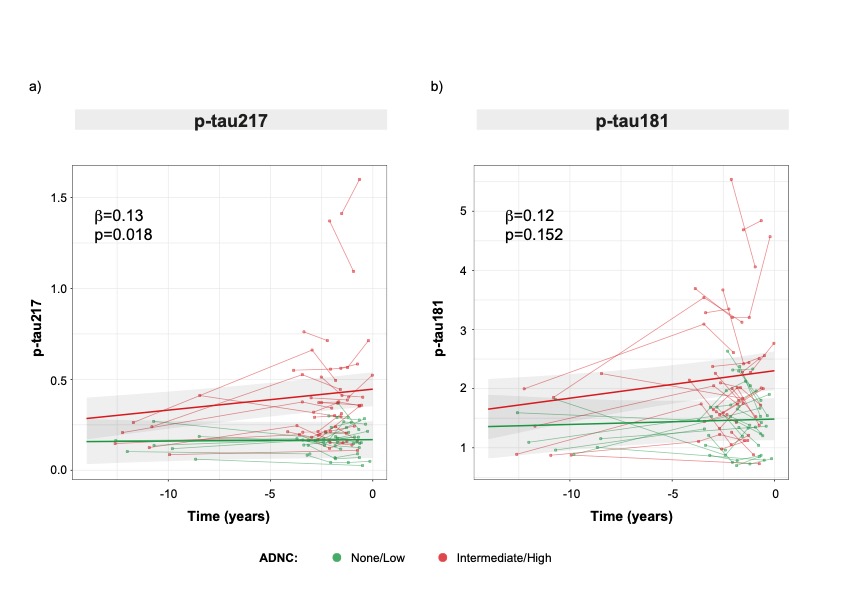

**Supplementary Figure 10** **Associations between longitudinal changes of plasma biomarkers and presence of ADNC at death**

Bold lines represent mean longitudinal changes of plasma p-tau217 (a) and plasma p-tau181 (b) by ADNC groups at death. Linear mixed effect models were used to derive these associations in independent models including: age at baseline, and sex as covariates using and random intercepts and fixed time-slopes. ADNC was dichotomized as negative (none/low) or positive (intermediate/high). ADNC*time interaction standardized betas and p-values are shown in the figure.

Abbreviations: ADNC, Alzheimer’s disease neuropathologic change; p-tau, phosphorylated tau.
